# Approaches in Analyzing Predictors of Trial Failure: A Scoping Review and Meta-epidemiological study

**DOI:** 10.1101/2025.08.08.25333296

**Authors:** Aleksa Jovanovic, Stojan Gavric, Fabio Dennstädt, Nikola Cihoric

## Abstract

**Importance:** Although there are numerous studies exploring predictors of clinical trial failure, there is a lack of structured knowledge of the methodological nuances of published studies in this field.

**Objective:** We performed a scoping review with the aim of exploring the methodological approaches in analyzing predictors of clinical trial failure.

**Evidence Review:** The Ovid Medline and Embase databases were systematically searched from inception to December 13, 2024, for studies employing frequentist statistics or machine learning (ML) approaches to assess predictors of trial failure across multiple clinical trials. A generalized linear model (GLM) was employed to assess the impact of methodological variations on reported failure proportions. To estimate the effects of the predictors included in the model on failure proportions, odds ratios (OR) with 95% confidence interval (95% CI) were calculated from model coefficients.

**Findings:** The literature search identified 17,961 records, 81 of which were included in the review. Most of the studies used Clinicaltrials.gov data (73 studies, 90.1%).

Frequentist statistics were used to analyze predictors of trial failure in 73 studies (90.1%), and remaining 8 studies employed ML techniques (9.9%). The GLM demonstrated that methodological factors explain 27.5% of the observed variability in failure proportions. Studies including both completed and ongoing status when calculating failure proportion had lower odds of failure compared to those just including completed status (OR = 0.44, 95% CI: 0.29–0.67, p < 0.001).

**Conclusions and Relevance:** There has been a recent expansion of ML approaches, potentially signaling the beginning of a paradigm shift. Methodological variations account for a significant amount of variation in failure proportion, signaling the need for adoption of standardized definitions of failure and calculation approach.

**Key Points:** *Question:* What are the methodological specificities of studies exploring predictors of clinical trial failure?

**Findings:** The choice of denominator and of included study type significantly influenced failure proportions. The use of machine learning to assess predictors of clinical trial failure is an emerging approach.

**Meaning:** There is a need for adoption of standardized definitions of trial failure and non- failure to have meaningful comparisons.

## Introduction

Contrary to popular belief, clinical trials with negative results should not be considered failed, since they contribute to the scientific knowledge base^1^. Furthermore, refusal to publish these negative results leads to publication bias which can later have negative implications on more comprehensive knowledge synthesis such as meta-analyses^2, 3^. On the other hand, clinical trials that do not meet recruitment goals or ensure adequate follow-up for endpoint evaluation lead to a loss of opportunities for expanding the knowledge base and can be considered failed. These prematurely terminated trials also have other direct and indirect consequences on the healthcare and scientific ecosystem, such as resource depletion and loss of motivation to reevaluate the same disease or interventions^4^.

Understanding the predictors of clinical trial failure is important as it can open opportunities for its prevention. Although there are numerous studies exploring trial failure predictors, there is a lack of structured knowledge of the methodological nuances of published studies in this field. Therefore, we performed a scoping review with the aim of evaluating the methodological landscape of studies exploring predictors of clinical trial failure.

## Methods

### Information sources and search strategies

We performed a scoping review according to the Preferred Reporting Items for Systematic Reviews and Meta-Analyses Extension for Scoping Reviews (PRISMA-ScR) Guidelines^5^ (eTable 1). The Ovid Medline and Embase databases were searched from inception to December 13, 2024. The complete search strategy for each of the databases is provided in eTables 2 and 3. An e-mail was sent to the corresponding author when the full text of a study fitting the inclusion criteria was unavailable.

### Eligibility criteria

Studies were included if they employed frequentist statistics or machine learning (ML) approaches to systematically assess predictors of trial failure (premature termination) across multiple clinical trials. We included cross-sectional, case-control, and modelling studies performing retrospective analyses of clinical trial registry data. No restrictions were placed on therapeutic area, demographics, publication date, language or type. Research articles, letters to the editor and conference abstracts all were deemed eligible if they fit the scope of the review. In the case of overlapping publications from the same research group, the most comprehensive report was included.

Studies were excluded if they: 1) presented predictor distributions without statistical analysis; 2) summarized existing literature without analysis; 3) analyzed predictors of trial failure due to isolated reasons such as failed accrual; 4 ) examined predictors non-systematically across only 1-2 trials; 5) were individual trial failure reports; 6) studied animal or cell models.

### Study selection

Recent research has shown that large language models (LLMs) can achieve high sensitivity in screening scientific publications for systematic reviews, with sensitivity rates of over 97% for certain models shown^6, 7^. The study selection process utilized the claude-3-5-sonnet-20241022 over Anthropic API LLM for initial screening of titles and abstracts. We employed a two-step approach, with an additional human check in step two, achieving 100% sensitivity and ∼84% specificity in each of the steps (eFigure 1). Details on the validation process and prompts used in the screening are presented in eAppendices 1 and 2, respectively. After screening titles and abstracts, the full text of papers assessed as relevant was obtained and assessed independently by two reviewers (A.J. and S.G.). The disagreements in this phase were resolved by consensus after involving the third reviewer (N.C).

**Figure 1.**
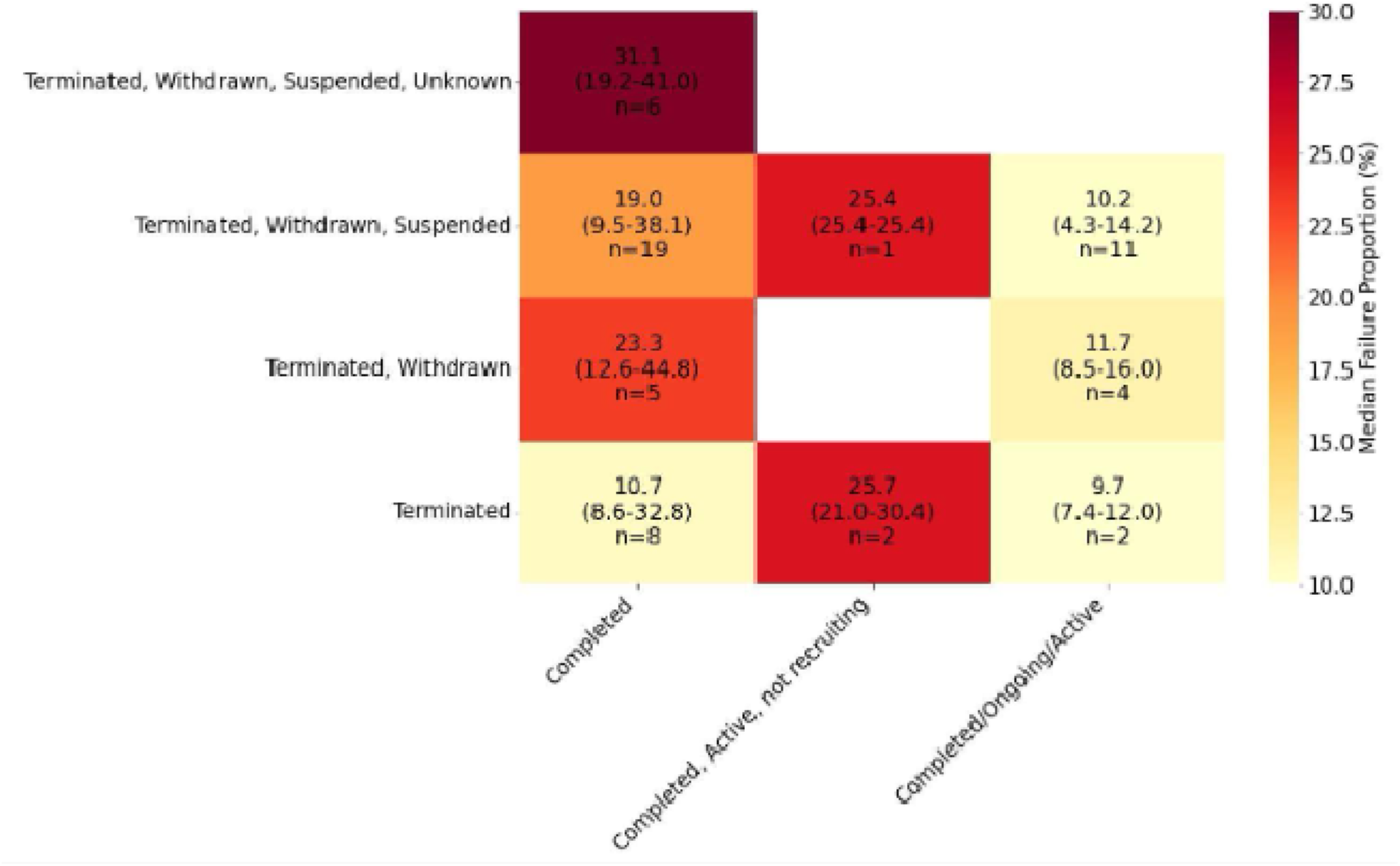
Failure proportion distribution by trial failure and non-failure definitions, median (range)

### Data charting and data items

A data charting form was developed jointly by two reviewers (A.J. and S.G.), who independently charted data. The included studies were assessed for the study meta- data (authors, title, year of publication), presence of results reporting and/or non-publication analysis, data source, therapeutic area, number of trials analyzed, eligibility criteria (study type, phase, demographics, trial location restrictions, etc.), failure definitions, non-failure definitions (i.e. the categories used as the denominator for computation of failure proportion), proportion of failed studies based on the definitions used, failure predictors assessed, distribution of failure reasons, statistical approach used for assessing failure predictors, statistically significant failure predictors in studies employing frequentist statistics, and methodological characteristics of ML studies (the types of structured and unstructured features used, feature engineering techniques, ML algorithms, evaluation metrics results, comparison to traditional methods, interpretability methods employed).

For studies using Clinicaltrials.gov datasets, the combined statuses of recruiting, enrolling by invitation, and active, not recruiting were categorized as active, while ongoing categorization was employed for these three statuses combined with the not yet recruiting status. When studies didn’t explicitly state which Clinicaltrials.gov definitions were used for failure proportion calculation, we made assumptions based on their methods and results section. In rare instances where a study used a different number of studies for predictor analysis and failure proportion calculation, we used the number related to failure proportions to maintain consistency.

During extraction of frequentist statistical methods, chi-square was extracted if it was the sole analysis conducted to explore associations between trial-level factors and trial failure, but wasn’t extracted when performed as a supplementary analysis to another, more sophisticated analyses (i.e. regression model). The exception was when the authors explicitly stated that the statistical significance of chi-square test was used to select variables for inclusion in the multivariable model.

### Synthesis of results

Therapeutic areas were grouped into broader categories (e.g. neuro-oncology was grouped into oncology category) when presenting descriptives. Descriptive statistics were used to calculate summary statistics of key characteristics of included studies.

A matrix of failure proportion distribution according to combinations of failure and non-failure definitions was computed for studies utilizing data from Clinicaltrials.gov (eNote 3). The analysis was repeated in two subsets of studies including only: a) interventional studies; b) randomized controlled trials (RCTs).

### Statistical analysis

Supplementary to our scoping review, we employed a generalized linear model (GLM) to quantify the extent to which methodological factors contribute to observed differences in failure proportions in studies analyzing different therapeutic areas. Therapeutic area couldn’t be incorporated as a variable in the model since the analyzed studies shared a common data source (Clinicaltrials.gov), which meant that certain trials would be counted multiple times across different studies, while others would appear only once. We avoided this bias by stratifying studies based on the therapeutic area and then selecting the most representative ones from each subset. A detailed explanation on the selection process of studies for the GLM, variable categorization and model selection are presented in eAppendix 4.

To estimate the effects of the predictors included in the model on failure proportions, odds ratios (OR) with 95% confidence interval (95% CI) were calculated from model coefficients. Significance was set at p<0.05. Analyses were conducted in R (version 4.3.1).

## Results

### Study selection

The literature search identified 17,961 records from Ovid Medline and Embase databases. After duplicate removal, the title and abstract screening phase was performed in multiple stages (eFigure 2), leading to 111 papers being assessed for eligibility, of which 93 were full papers, and 18 were conference abstracts. After assessing full manuscripts and conference abstracts for relevance, 81 studies^8–88^ were finally chosen for inclusion (eFigure 2). Of these, 7 were reports from a conference, and for a single paper the full text couldn’t be obtained even after contacting the authors. Due to ambiguity of failure and non-failure definitions and limited comprehensiveness of the data available in these 8 abstracts, they were excluded from failure classification analyses.

**Figure 2.**
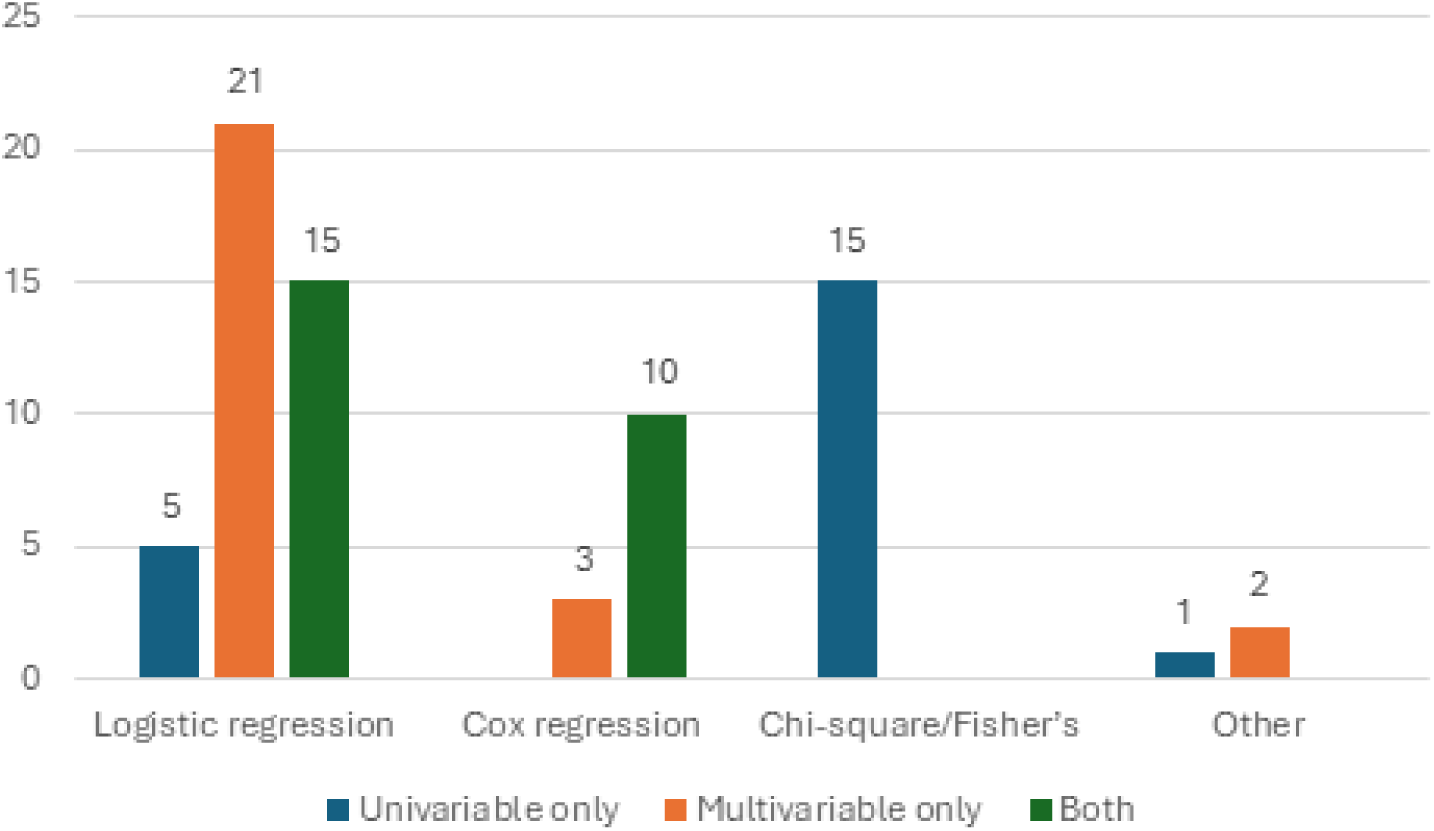
Distribution of frequentist statistics methodologies employed for predictor analysis.

### Characteristics of included studies

Most of the 81 included studies used Clinicaltrials.gov data (73 studies, 90.1%) (Table 1). More than a third of included papers (32, 39.5%) assessed predictors for non-publication in addition to predictors of premature termination, while a minority (11, 13.6%) assessed the predictors of results reporting.

**Table 1.** Distribution of characteristics of included studies.

| Study characteristic | All studies (N=81)<br>Studies, No. (%) | Full text studies<br>analysing Ct.gov<br>(N=66)<br>Studies, No. (%) |
| --- | --- | --- |
| Data source |  |  |
| Ct.gov | 73 (90.1) | 66 (100.0) |
| Other national databases | 5 (6.2) | 0 (0.0) |
| EudraCT | 2 (2.5) | 0 (0.0) |
| Medline | 1 (1.2) | 0 (0.0) |
| Topic |  |  |
| Oncology | 21 (25.9) | 19 (28.8) |
| Surgery | 13 (16.1) | 13 (19.7) |
| Internal medicine | 9 (11.1) | 4 (6.1) |
| No restriction | 15 (18.5) | 9 (13.6) |
| Neurology | 5 (6.2) | 4 (6.1) |
| Obstetrics and gynecology | 4 (4.9) | 4 (6.1) |
| Pediatrics | 4 (4.9) | 3 (4.5) |
| Psychiatry | 3 (3.7) | 3 (4.5) |
| Other | 7 (8.7) | 7 (10.6) |
| Study type included |  |  |
| Any | 16 (19.7) | 12 (18.2) |
| Interventional only | 43 (53.1) | 35 (53.0) |
| RCT only | 22 (27.2) | 19 (28.8) |
| Failed study definition | NA |  |
| Terminated, Withdrawn or Suspended |  | 31 (47.0) |
| Terminated |  | 12 (18.2) |
| Terminated or Withdrawn |  | 10 (15.1) |
| Terminated, Withdrawn, Suspended or Unknown |  | 6 (9.1) |
| Other 6 definitions |  | 7 (10.6) |
| Non-failed study definition | NA |  |
| Completed |  | 38 (57.6) |
| Completed, Ongoing*, Unknown |  | 12 (18.2) |
| Other 12 definitions |  | 16 (24.2) |

\*Ongoing entails Recruiting, Enrolling by invitation, Active, not recruiting, and Not yet recruiting statuses

The most frequently analyzed major therapeutic area was oncology (21, 25.9%), followed by studies analyzing the dataset comprehensively, without restrictions on the therapeutic area or demographics (15, 18.5%) (Table 1). Seven of the studies (8.7%) each analyzed a unique therapeutic area (Table 1).

Most studies excluded observational studies from the analysis (65, 80.2%), while about a quarter of the total studies (22, 27.2%) focused exclusively on RCTs, also excluding non-randomized clinical trials (Table 1). The median number of included trials per study was 746 (range 54-284,644). The median failure proportion of all included studies was 18%, ranging from 3% in a study analyzing 312 drug clinical trials registered in China, to 51.8% in a study analyzing 85 cervical cancer and precancer trials registered at Clinicaltrials.gov (eTable 4).

### Clinical trial failure classifications

There were 10 distinct clinical trial failure definitions among the 66 full text papers analyzing Clinicaltrials.gov (eTable 4), with almost half of the studies defined failure as terminated, withdrawn or suspended trial status (31 studies, 47.0%) (Table 1). When considering non-failure definitions, i.e. study statuses beside failed that were included in the denominator when calculating the failure proportion, the diversity was even greater. There were 14 distinct definitions among the 65 papers (eTable 4), with more than half of the papers including only the completed status (38 studies, 57.6%) (Table 1).

Failure proportions varied depending on the combination of failure and non-failure definitions used (Figure 1). After merging categories and discarding unique categories (eAppendix 3), 57 studies with 4 distinct failure and 3 distinct non-failure definitions for a total of 9 combinations were left in the analysis. The lowest failure proportion was observed among 2 studies with terminated (failed) and completed, ongoing/active (non-failed) definitions combination (median 9.7 range 4.3-14.2), while the highest was observed when trial failure was defined as terminated, withdrawn, suspended, or unknown, and non-failure was defined as completed (median 31.1, range 19.2-41.0). The distribution of failure proportions according to failure and non-failure definitions in studies excluding observational trials and studies including only RCTs are shown in eFigures 3 and 4, respectively.

### Influence of methodological choices on observed failure proportions

The beta-binomial model (AIC = 237.06) outperformed the null model (AIC = 250.66) and explained 27.5% of the total variability in failure proportions (deviance reduction from 100.26 to 72.66), indicating that methodological factors account for about a quarter of observed differences, while the remaining variability likely reflects unmeasured factors such as therapeutic area.

Non-failure definition and study type were statistically significant predictors of failure proportions in the beta-binomial model. Studies including completed/ongoing when calculating failure proportion had lower odds of failure compared to those just including completed status as non-failure (OR = 0.44, 95% CI: 0.29–0.67, p < 0.001). Studies including strictly randomized trials had higher odds of failure than studies including both randomized and non-randomized trials (OR = 1.89, 95% CI: 1.12–3.19, p = 0.016), while studies including both observational and interventional studies had lower odds (OR = 0.53, 95% CI: 0.34–0.85, p = 0.007). The failure definitions and phase restrictions weren’t statistically significant predictors of failure proportions.

### Statistical approaches in assessing failure predictors

All but eight studies identified in the review (73 studies, 90.1%) used frequentist statistics to assess predictors of trial failure. Among these, logistic regression was most frequently used (41 studies, 56.2%), followed by chi-square/Fisher’s exact test (15 studies, 20.8%), and Cox regression analysis (13 studies, 18.1%). One study employed multivariable Poisson regression, one employed GLM, one claimed that “relative risks” were calculated without stating which test was used, and one conference abstract stated that “hazard ratio analysis” was performed, but results presented ORs, prompting us to exclude the study from the distribution analysis due to ambiguity. Almost three quarters (52 studies, 72.2%) of studies employing frequentist statistics employed a multivariable analysis controlling for confounding, with two employing multivariable logistic regression chosing the predictors to include in the multivariable model based on the significance obtained after performing chi-square test (Figure 2).

Eight studies employed ML approaches to analyze predictors of trial failure, with the first study published in 2019, and half published in 2023. Approaches ranged from conventional statistical learning to modern neural architectures (Table 2). Four studies used established techniques such as LASSO, Random Forest, gradient boosting, while others employed more specialized approaches: Latent Dirichlet Allocation (LDA) for topic extraction from trial protocols by using probabilistic topic modeling, neural networks for survival analysis, and transformer-based language models for text processing.

**Table 2.**
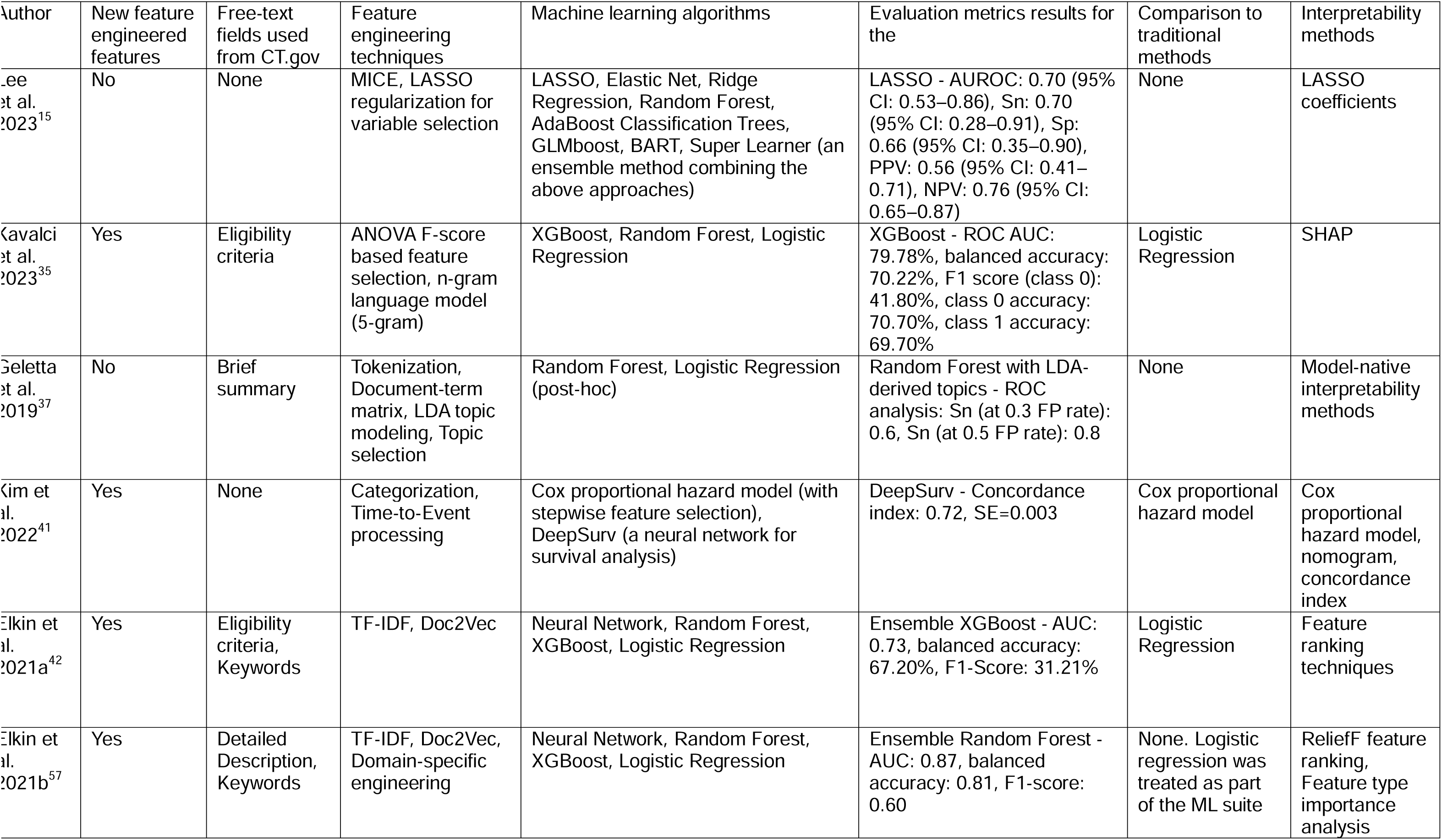
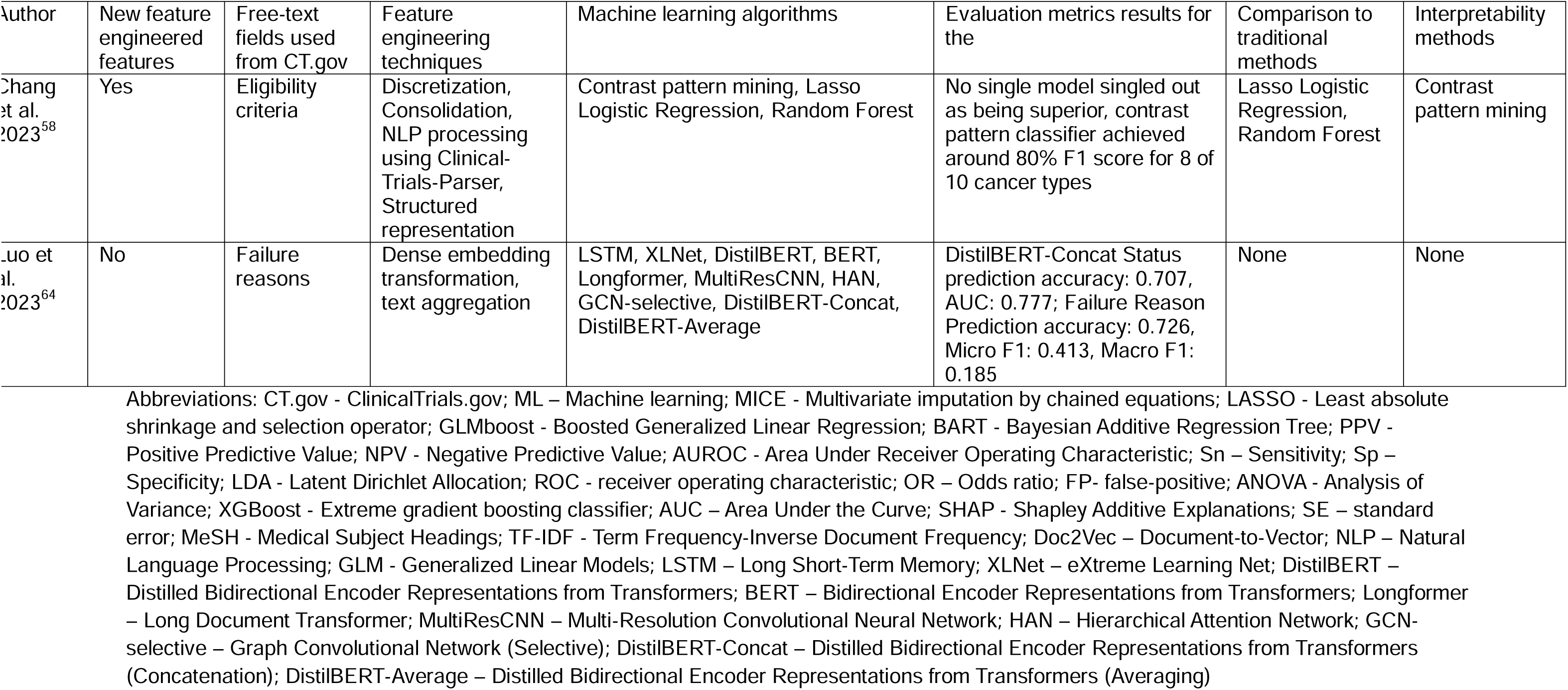
Methodological characteristics of machine learning papers.

### Trial failure predictors

Table 3 shows the distribution of results of multivariable analyses analyzing predictors of trial failure. The statistically significant findings from multivariable analysis of failure predictors from included studies are presented in eTable 5, with a detailed summary provided in eAppendix 5.

**Table 3.** Predictors of trial failure.

| Predictor | Studies analyzing the predictor<br>Studies, No. (%) | Failure risk factor<br>Studies, No. (%) | Protective of failure<br>Studies, No. (%) | Non-significant<br>Studies, No. (%) |
| --- | --- | --- | --- | --- |
| Enrollment | 25 (100) | 18 (72) | 0 (0) | 7 (28) |
| Anticipated accrual | 8 (100) | 1 (12.5) | 2 (25) | 5 (62.5) |
| Location (US) | 18 (100) | 10 (55.6) | 0 (0) | 8 (44.4) |
| Higher number of participating countries | 3 (100) | 1 (33.3) | 0 (0) | 2 (66.7) |
| Higher number of participating centers | 21 (100) | 5 (23.8) | 3 (14.3) | 13 (61.9) |
| Blinding | 23 (100) | 3 (13) | 3 (13) | 17 (73.9) |
| Randomization | 16 (100) | 2 (12.5) | 0 (0) | 14 (87.5) |
| Presence of a data monitoring committee | 3 (100) | 1 (33.3) | 1 (33.3) | 1 (33.3) |

## Discussion

Our scoping review revealed significant methodological differences in defining failure and selecting trial statuses for denominators, contributing to wide variation in reported failure proportions (4.3%–46.1%). Our model attributed about 27.5% of this variance to these definitions and included study designs.

The extremes of the failure proportions illustrate how denominator selection impacts its value. The five studies with the highest failure proportions (40.1%–46.1%) all used restrictive denominators (i.e., only completed trials as non-failures) but differed in failure definitions (all included terminated and withdrawn, some additionally included suspended and/or unknown)^11, 15, 64, 80, 85^. Additionally, some of these studies had important specificities further inflating the failure proportions: counting only studies with published results as completed^80^; not including all completed trials because of implementation of a 1:1 case-control design^64^; or categorizing studies completed with <85% of targeted sample size as failed^15^. The five studies had widely different scope (urology, therapy-related, cancer trials in older adults, neuro-oncology, and extremity fracture), indicating that the investigated therapeutic area isn’t the only driver of differences between failure proportions.

Furthermore, the studies reporting the lowest failure proportions (4.3% and 5.6%) included all 8 Clinicaltrials.gov statuses in the denominator, thereby diluting the failure proportions^36, 54^. Including ongoing trials when reporting failure proportions underestimates the true failure proportion since some of these trials will ultimately fail, but none of them are counted as failed since, at prevalence day, their outcomes are still unknown. The beta-binomial model showed that studies including ongoing or active trials in their denominators reported 53% lower odds of failure compared to those excluding these trials.

Analyzing predictors of failure while including ongoing studies can lead to misleading implications, since these studies have systematically different trial characteristics compared to completed ones^89^. Similarly, a common practice of counting suspended trials as failed should be challenged, since these trials, by definition, could restart and complete successfully^90^. We propose standardizing definitions for future research, recommending categorizing terminated and withdrawn studies as failed and only completed ones as non-failed. This approach would provide more accurate measurements of prevalence and predictors of trial failure by eliminating the confounding of studies that still have the potential to be completed either successfully or unsuccessfully.

Another methodological difference influencing the observed failure proportions is the type of studies included in the analysis. This is in line with the association observed in several studies that randomization is associated with trial failure^51, 68, 75, 88^. Analyses restricted to RCTs showed 90% higher failure odds compared to those including both randomized and non-randomized interventional trials. This suggests might RCTs face greater completion challenges due to their more rigorous methodological requirements, although the findings from the multivariable analyses for randomization as the risk factor for failure were not convincing^9, 12, 13, 16, 22, 29, 30, 38, 41, 50, 52–54, 65, 79, 80^. On the other hand, studies including observational trials showed lower failure odds (OR=0.52, 95% CI: 0.33-0.82), consistent with the generally simpler execution of observational studies^27, 51, 56, 72^.

In our review, we identified eight studies that have employed ML techniques to analyze predictors of trial failure^15, 35, 37, 41, 42, 57, 58, 64^. This novel approach has several advantages over the traditional statistical methods. First, it can handle a much larger number of features, and collinearity and non-linearity are not issues as in traditional statistical models^91^. Second, the ML approach has the option to automatically detect feature interactions. Third, it has the ability to handle unstructured data (e.g., trial protocols, eligibility criteria)^92^. Additionally, by employing interpretability techniques, it can rank the relative importance of different predictors in the model’s decision. As opposed to traditional statistics’ summary statistics, these interpretability techniques can be implemented to give a risk profile based on the relative importance of different features integrated into the model on a study-by-study basis^93^. This is important for clinical trials’ stakeholders, as it provides transparency needed to justify potential protocol alterations. Further development and implementation of these models could lead to reducing clinical trial failure proportions.

Our study has several limitations. First, although we implemented a multi-iterative LLM-assisted screening approach and validated it showing 100% sensitivity, the possibility remains that some relevant studies were missed. However, due to the vast number of correctly identified and included studies in the review, and the absence of false negatives in categories 2 or 3 in the more complex abstract screening step, we deem it highly unlikely. Second, our analysis of methodological factors affecting failure proportions was limited by the potential overlap in trials, forcing us to exclude several studies with larger sample sizes from modelling. This also prevented us from including therapeutic area in the model to provide an estimate on how much of the trial failure proportion variation can be attributed to it. Finally, the specific impact of many rare non-failure definitions could not be analyzed, due to lack of studies, however, our model encompassed the most important and the most frequent categories.

## Conclusion

Our findings show the limitations of comparing failure proportions across therapeutic areas by comparing calculated proportions across studies. A need exists for adoption of standardized definitions of trial failure and non-failure for more accurate comparisons. Finally, the recent expansion of ML models for predicting trial failure shows a promising prospect for reducing clinical trial failure in the future.

## Data Availability

The data sets used and/or analyzed during the study are available from the corresponding author on request.

## Ethical approval

The study involves no human subjects and requires no ethical approval.

## Consent for publication

Not applicable.

## CRediT authorship contribution statement

**Aleksa Jovanovic:** Writing – original draft, Writing – review & editing, Visualization, Validation, Methodology, Investigation, Formal analysis, Data curation, Conceptualization. **Stojan Gavric:** Writing – review & editing, Methodology, Visualization, Formal analysis, Data curation, Software. **Fabio Dennstädt:** Writing – review & editing, Visualization, Data curation. **Nikola Cihoric:** Writing – review & editing, Supervision, Project administration, Conceptualization.

**Declaration of generative AI and AI-assisted technologies in the writing process** During the preparation of this work the authors used claude-3-5-sonnet-20241022 over Anthropic API LLM for initial screening of titles and abstracts. After using this tool, the author(s) reviewed and edited the content as needed, including performing validation as outlined in the manuscript and supplementary material and take full responsibility for the content of the publication.

