## Supplementary material for "Approaches in Analyzing Predictors of Trial Failure: A Scoping Review and Meta-epidemiological study"

**Supplemental Online Content**

**eTable 1** Preferred Reporting Items for Systematic reviews and Meta-Analyses extension for Scoping Reviews (PRISMA-ScR) Checklist

**eTable 2** Embase search strategy

**eTable 3** Ovid Medline Search Strategy

**eAppendix 1** Prompt engineering and screening validation process

**eAppendix 2** Prompts used for AI title and abstract screening

**eFigure 1** Confusion matrices for the performance of Phase 1 (title screening) and Phase 2 (chain-of-thought abstract screening)

**eAppendix 3** Data cleaning performed prior to computing the failure proportion distribution by trial failure and non-failure definitions matrix

**eAppendix 4** Generalized linear model methodology

**eFigure 2** PRISMA Flowchart of the included studies

**eTable 4** Characteristics of included studies

**eFigure 3** Failure proportion distribution by trial failure and non-failure definitions, studies including interventional studies only

**eFigure 4** Failure proportion distribution by trial failure and non-failure definitions, studies including randomized controlled trials only

**eTable 5** Statistically significant predictors of study failure in the multivariable analysis

**eTable 1 Preferred Reporting Items for Systematic reviews and Meta-Analyses extension for Scoping Reviews (PRISMA-ScR) Checklist**

| **SECTION** | **ITEM** | **PRISMA-ScR CHECKLIST ITEM** | **REPORTED ON PAGE #** |
| --- | --- | --- | --- |
| **TITLE** | | | |
| Title | 1 | Identify the report as a scoping review. | 1 |
| **ABSTRACT** | | | |
| Structured summary | 2 | Provide a structured summary that includes (as applicable): background, objectives, eligibility criteria, sources of evidence, charting methods, results, and conclusions that relate to the review questions and objectives. | 2-3 |
| **INTRODUCTION** | | | |
| Rationale | 3 | Describe the rationale for the review in the context of what is already known. Explain why the review questions/objectives lend themselves to a scoping review approach. | 4 |
| Objectives | 4 | Provide an explicit statement of the questions and objectives being addressed with reference to their key elements (e.g., population or participants, concepts, and context) or other relevant key elements used to conceptualize the review questions and/or objectives. | 4 |
| **METHODS** | | | |
| Protocol and registration | 5 | Indicate whether a review protocol exists; state if and where it can be accessed (e.g., a Web address); and if available, provide registration information, including the registration number. | 4 |
| Eligibility criteria | 6 | Specify characteristics of the sources of evidence used as eligibility criteria (e.g., years considered, language, and publication status), and provide a rationale. | 5 |
| Information sources* | 7 | Describe all information sources in the search (e.g., databases with dates of coverage and contact with authors to identify additional sources), as well as the date the most recent search was executed. | 4-5 |
| Search | 8 | Present the full electronic search strategy for at least 1 database, including any limits used, such that it could be repeated. | eTables 1 and 2 |
| Selection of sources of evidence† | 9 | State the process for selecting sources of evidence (i.e., screening and eligibility) included in the scoping review. | 5-6 |
| Data charting process‡ | 10 | Describe the methods of charting data from the included sources of evidence (e.g., calibrated forms or forms that have been tested by the team before their use, and whether data charting was done independently or in duplicate) and any processes for obtaining and confirming data from investigators. | 6 |
| Data items | 11 | List and define all variables for which data were sought and any assumptions and simplifications made. | 6-7 |
| Critical appraisal of individual sources of evidence§ | 12 | If done, provide a rationale for conducting a critical appraisal of included sources of evidence; describe the methods used and how this information was used in any data synthesis (if appropriate). | N/A |
| Synthesis of results | 13 | Describe the methods of handling and summarizing the data that were charted. | 7 |
| **RESULTS** | | | |
| Selection of sources of evidence | 14 | Give numbers of sources of evidence screened, assessed for eligibility, and included in the review, with reasons for exclusions at each stage, ideally using a flow diagram. | eFigure 2 |
| Characteristics of sources of evidence | 15 | For each source of evidence, present characteristics for which data were charted and provide the citations. | eTable 4 |
| Critical appraisal within sources of evidence | 16 | If done, present data on critical appraisal of included sources of evidence (see item 12). | N/A |
| Results of individual sources of evidence | 17 | For each included source of evidence, present the relevant data that were charted that relate to the review questions and objectives. | Table 1, Table 2, Table 3, Figure 1, Figure 2 |
| Synthesis of results | 18 | Summarize and/or present the charting results as they relate to the review questions and objectives. | 9-12 |
| **DISCUSSION** | | | |
| Summary of evidence | 19 | Summarize the main results (including an overview of concepts, themes, and types of evidence available), link to the review questions and objectives, and consider the relevance to key groups. | 12-15 |
| Limitations | 20 | Discuss the limitations of the scoping review process. | 15 |
| Conclusions | 21 | Provide a general interpretation of the results with respect to the review questions and objectives, as well as potential implications and/or next steps. | 15-16 |
| **FUNDING** | | | |
| Funding | 22 | Describe sources of funding for the included sources of evidence, as well as sources of funding for the scoping review. Describe the role of the funders of the scoping review. | 16 |

JBI = Joanna Briggs Institute; PRISMA-ScR = Preferred Reporting Items for Systematic reviews and Meta-Analyses extension for Scoping Reviews.

* Where *sources of evidence* (see second footnote) are compiled from, such as bibliographic databases, social media platforms, and Web sites.

† A more inclusive/heterogeneous term used to account for the different types of evidence or data sources (e.g., quantitative and/or qualitative research, expert opinion, and policy documents) that may be eligible in a scoping review as opposed to only studies. This is not to be confused with *information sources* (see first footnote).

‡ The frameworks by Arksey and O’Malley (6) and Levac and colleagues (7) and the JBI guidance (4, 5) refer to the process of data extraction in a scoping review as data charting*.*

§ The process of systematically examining research evidence to assess its validity, results, and relevance before using it to inform a decision. This term is used for items 12 and 19 instead of "risk of bias" (which is more applicable to systematic reviews of interventions) to include and acknowledge the various sources of evidence that may be used in a scoping review (e.g., quantitative and/or qualitative research, expert opinion, and policy document).

*From:* Tricco AC, Lillie E, Zarin W, O'Brien KK, Colquhoun H, Levac D, et al. PRISMA Extension for Scoping Reviews (PRISMAScR): Checklist and Explanation. Ann Intern Med. 2018;169:467–473. [doi: 10.7326/M18-0850](http://annals.org/aim/fullarticle/2700389/prisma-extension-scoping-reviews-prisma-scr-checklist-explanation).

**eTable 2 Embase search strategy (search conducted on 13.12.2024.)**

| # | Searches | Results |
| --- | --- | --- |
| 1 | ((complet* OR unsuccess* OR fail* OR incomplet* OR noncomplet* OR discontinu* OR termin* OR suspen* OR withdraw* OR halt*):ti AND (trial* OR "clinical study" OR "clinical studies" OR RCTs OR RCT OR “clinical trial*” OR premature* OR early):ti) OR 'early termination of clinical trials'/exp | 30,891 |
| 2 | 'clinical trial'/exp OR 'trial level':ab,ti OR 'research design'/exp OR 'case control study'/exp OR 'cross-sectional study'/exp OR 'retrospective study'/exp OR 'systematic review'/de OR 'meta analysis'/de OR cross-sectional:ab,ti OR case-control:ab,ti OR systematic review:ab,ti OR meta-analysis:ab,ti | 1,440,353 |
| 3 | 'database'/exp OR 'registry'/exp OR database*:ab,ti OR dataset*:ab,ti OR data set:ab,ti OR registry*:ab,ti OR ClinicalTrials.gov:ab,ti OR ct.gov:ab,ti OR www.clinicaltrials.gov:ab,ti OR ClinicalTrials:ab,ti OR ICTRP:ab,ti OR EUCTR:ab,ti OR ISRCTN:ab,ti | 802,376 |
| 4 | 'machine learning'/exp OR 'natural language processing'/exp OR 'artificial intelligence'/exp OR 'algorithm'/exp OR 'neural network'/exp OR data mining:ab,ti OR secondary data analy*:ab,ti OR machine learning:ab,ti OR neural network*:ab,ti OR decision tree:ab,ti OR random forest:ab,ti OR big data:ab,ti OR deep learning:ab,ti OR support vector machine*:ab,ti OR contrast mining:ab,ti OR gradient boost*:ab,ti OR xgboost:ab,ti OR bert:ab,ti OR transformer*:ab,ti OR bayesian network*:ab,ti OR ensemble learn*:ab,ti OR interpretab*:ab,ti | 66,504 |
| 5 | 'logistic model'/exp OR 'regression analysis'/exp OR 'multivariate analysis'/exp OR 'risk factor'/exp OR 'statistical model'/exp OR 'area under curve'/exp OR 'eligibility determination'/exp OR 'probability'/exp OR predictive model*:ab,ti OR risk model*:ab,ti OR nomogram*:ab,ti OR model validat*:ab,ti OR model calibrat*:ab,ti OR multivar* analy*:ab,ti OR logistic regression:ab,ti OR odds ratio:ab,ti OR risk factor*:ab,ti OR regression analy*:ab,ti OR proportional hazard*:ab,ti OR area under curve:ab,ti OR receiver operator characteristic:ab,ti | 362,684 |
| 6 | #2 OR #3 OR #4 OR #5 | 2,551,474 |
| 7 | #1 AND #6 | 5,189 |

**eTable 3 Ovid Medline Search Strategy (search conducted on 13.12.2024.)**

| # | Searches | Results |
| --- | --- | --- |
| 1 | ((complet* OR unsuccess* OR fail* OR incomplet* OR noncomplet* OR discontinu* OR termin* OR suspen* OR withdraw* OR halt*).ti AND (trial* OR "clinical study" OR "clinical studies" OR RCT* “RCTs” OR “clinical trial*” OR premature* OR early).ti) OR exp "early termination of clinical trials"/ | 21886 |
| 2 | exp "clinical trial"/ OR exp "research design"/ OR exp "case-control studies"/ OR exp "cross-sectional studies"/ OR exp "retrospective studies"/ OR exp "systematic review"/ OR exp "meta-analysis"/ OR (cross-sectional OR case-control OR systematic review OR meta-analysis).ti,ab | 3974223 |
| 3 | exp "databases, factual"/ OR exp "registries"/ OR (database* OR dataset* OR data set OR registry* OR “ClinicalTrials.gov” OR “ct.gov” OR “[www.clinicaltrials.gov](http://www.clinicaltrials.gov)” OR ClinicalTrials OR ICTRP OR EUCTR OR ISRCTN).ti,ab | 1440823 |
| 4 | exp "machine learning"/ OR exp "natural language processing"/ OR exp "artificial intelligence"/ OR exp "algorithms"/ OR exp "neural networks (computer)"/ OR (data mining OR secondary data analy* OR machine learning OR neural network* OR decision tree OR random forest OR big data OR deep learning OR support vector machine* OR contrast mining OR gradient boost* OR xgboost OR bert OR transformer* OR bayesian network* OR ensemble learn* OR interpretab*).ti,ab | 693788 |
| 5 | exp "logistic models"/ OR exp "regression analysis"/ OR exp "multivariate analysis"/ OR exp "risk factors"/ OR exp "statistical models"/ OR exp "ROC curve"/ OR exp "eligibility determination"/ OR exp "probability"/ OR (predictive model* OR risk model* OR nomogram* OR model validat* OR model calibrat* OR multivar* analy* OR logistic regression OR odds ratio OR risk factor* OR regression analy* OR proportional hazard* OR area under curve OR receiver operator characteristic).ti,ab | 3151904 |
| 6 | 2 OR 3 OR 4 OR 5 | 7149858 |
| 7 | 1 AND 6 | 12772 |

**eAppendix 1 Prompt engineering and screening validation process**

The goal was to calibrate the large language model (LLM) to provide no false negative results, while minimizing clearly irrelevant studies required for human review. Several iterations of the prompt were made, with the LLM’s performance assessed after each iteration. Since most of the papers yielded by the search strategy are irrelevant to the review, the final iteration employed a two-step approach in order to increase efficiency:

a) First, the model was prompted to classify paper titles as either relevant or not relevant to clinical trial failure. This step was employed with the aim of filtering out the clearly irrelevant papers that are simple for the LLM to discern, therefore reducing the workload for the second, more demanding step. Here we chose to focus on the broader category of trial failure as opposed to focusing specifically on predictors on trial failure to avoid the LLM excluding studies due to insufficient data in the title. This resulted in the increase of the studies labeled as relevant not actually being relevant for inclusion in the review, therefore reducing the specificity metric (see below).
b) In the second step, the papers classified as relevant in the first step underwent a detailed chain-of-thought analysis based on detailed pre-defined criteria. The LLM assigned these papers into five categories (1: Clearly Irrelevant, 2: Probably Irrelevant, 3: Limited Generalizability, 4: Possibly Relevant, and 5: Highly Relevant) and was instructed to provide the rationale for its decision. The 5-category classification was employed to provide leeway for the LLM not to miss papers it might find ambiguous (2-4). This granular approach also facilitated human review by allowing increased focus on studies categorized as 4-5, while keeping the visibility of papers categorized as 2-3 (as opposed to losing the visibility in the case they were categorized as 1 alongside completely irrelevant papers).

We performed validation of both stages of the LLM classification by drawing a random sample of papers needed to achieve a 95% confidence interval (95% CI) with 5% margin of error (ME) and afterwards reviewing the accuracy of the model’s decisions in the samples by a human reviewer (A.J.). In both steps, positive cases were classified as papers that fit the scoping review inclusion criteria based on the abstract, while negative cases were classified as papers that didn’t fit the inclusion criteria. As stated previously, this led to most of the papers AI correctly categorized as relevant in the first step (concerning clinical trial failure in general) being labeled as false positive since they didn’t fit the inclusion criteria for the scoping review; and similarly for papers correctly classified into categories 2-4 in the second step.

No relevant papers classified as not relevant were found among the random sample of 375 papers analyzed (out of 14,773) in the first step or among the 325 analyzed (out of 2,120) in the second step, achieving 100% sensitivity in identifying papers that should be included in the review in both steps. The specificity in the first step was 84.6%, and 83.9% in the second step.

After the model validation was performed and the AI performance was deemed as appropriate, a reviewer (A.J.) examined the titles of papers in categories 2 and 3 (finding no relevant papers) and read detailed abstracts for categories 4 and 5.

**eAppendix 2 Prompts used for AI title and abstract screening**

**Final prompt used for title screening**

Role:user
Text:Classify this paper title for its relevance to clinical trial failure: Title: {title} Scoring criteria: - Score 1: the main topic of the paper is related to trial failure/trial non-completion or trial termination - Score 0: Paper discusses unrelated topics or is a standard RCT report

**Final prompt used for abstract screening**

Role: user

Text: You are a clinical trials research methodology expert tasked with classifying papers based on their relevance for inclusion in the scoping review identifying approaches in examining factors leading to clinical trial early termination. Your goal is to provide a single numerical classification (1-5) for each paper based on its title and abstract.

Here is the title of the paper you need to classify:

<title>

{{title_text}}

</title>

And here is the abstract:

<abstract>

{{abstract_text}}

</abstract>

Classification Scale:

**1. Clearly Irrelevant**

- Entirely unrelated to trial failure factors analysis

- Discusses treatment failure, discontinuation, power failure, or reporting completeness without linking to trial failure factors

- Covers tangential topics (e.g., enrollment barriers, gender disparities) without direct trial failure analysis

- Reports outcomes from a single terminated trial - Contains no analysis methodology for investigating trial failures

**2. Probably Irrelevant**

- Discusses trial failure without analyzing predictors of trial failure.

**3. Limited Generalizability**

- Investigates trial failure factors but lacks systematic methodology or analyzes few trials (1-5)

- Includes narrative reviews, exploratory reviews, qualitative studies, or editorials about trial failure factors without systematic methodology

- May summarize failure rates or reasons without comprehensive exploration of contributing factors

**4. Possibly Relevant**

- The study MUST analyze trial success or failure predictors

- If the study analyzes trial success or failure predictors it must employ systematic methodology on a large number of trials, but does so descriptively, without any sophisticated analysis performed

- Both of the previous criteria must be fulfilled to be considered possibly relevant. Do NOT consider it for possibly relevant if it is only a descriptive analysis without analysing predictive factors of trial failure.

**5. Highly Relevant**

- Employs robust systematic or data-driven approaches (e.g., retrospective analyses, predictive modeling, large dataset evaluations) to analyze factors contributing to clinical trial failure, termination, or non-completion.

- Focuses on analyzing specific operational, design, patient-related, or other contributing factors systematically linked to trial failure outcomes

Instructions:

1. Carefully read the title and abstract.

2. In <classification_analysis> tags, break down your thought process:

- Quote relevant parts of the title and abstract that inform your classification.

- Determine if the paper is about clinical trials early termination, non-completion or failure.

- Identify and count original analysis of predictors contributing to trial failure or discontinuation.

- Assess the comprehensiveness of the analysis, if present, and evaluate the methodology used.

- Consider any statistical tests or analyses related to trial discontinuation or failure, even if they're not the primary focus of the study.

- Explicitly consider arguments for each classification level (1-5), noting the strengths of each relevant category.

3. Based on your analysis, choose the most appropriate classification number (1-5).

4. Provide a brief explanation for your classification choice.

5. Output ONLY the classification number as your final response.

Example output structure:

<classification_analysis>

Relevant quotes:

"[Quote from title]"

"[Quote from abstract]"

The paper [does/does not] mention trial failure.

Predictors: [List and count analyzed trial success or failure predictors if any]

Methodology: [Describe methodology if mentioned]

Arguments for each classification:

1. [Argument for 1]

2. [Argument for 2]

3. [Argument for 3]

4. [Argument for 4]

5. [Argument for 5]

The strongest argument is for classification [X] because [reason].

</classification_analysis>

[Single digit classification number]

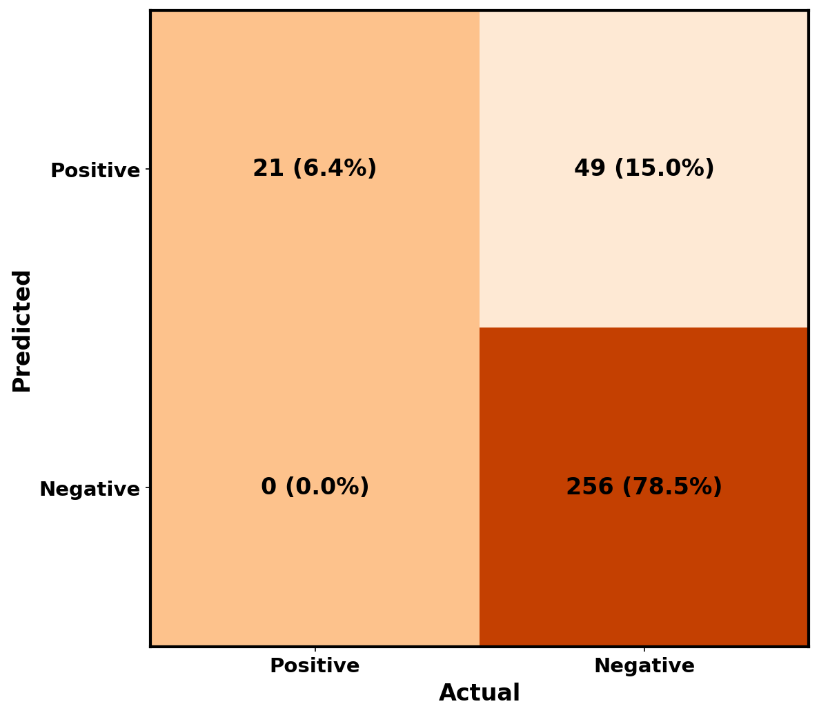

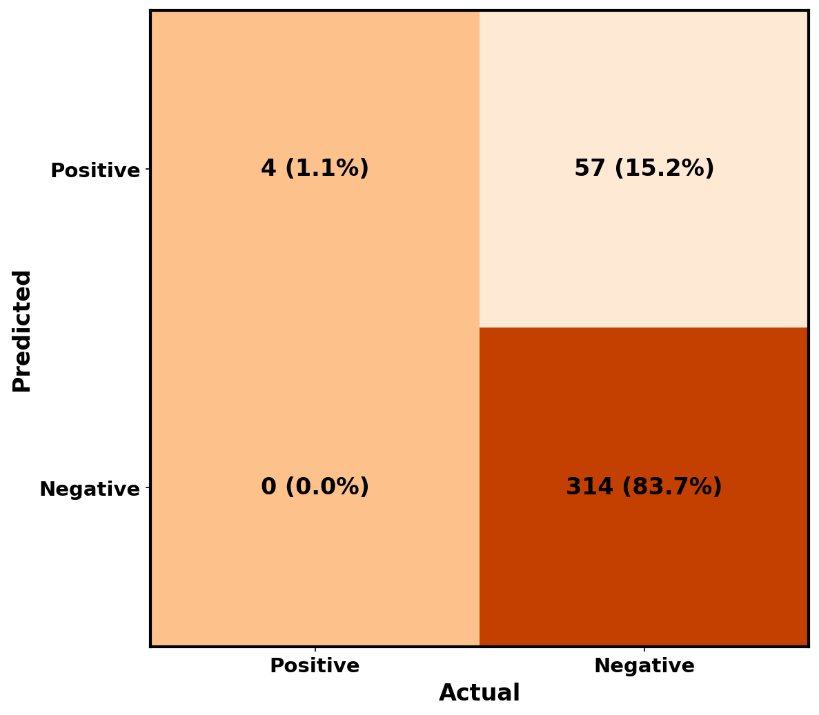

**eFigure 1 Confusion matrices for the performance of Phase 1 (title screening) (left) and Phase 2 (chain-of-thought abstract screening) (right)**

**eAppendix 3 Data cleaning performed prior to computing the failure proportion distribution by trial failure and non-failure definitions matrix**

Studies that employed unique definitions of failure and/or non-failure were excluded from this analysis, while some similar categories were merged.

**Removed**

1. Study ^35^ - removed because no data on failure proportion
2. Study ^15^ – removed because unique definition with 85% sample size AND also has terminated in the non-failed as well
3. Studies ^30, 36, 41^ - removed because they were all unique within their non failure categories, all had suspended in it, and it was not possible to group them together. Study ^86^ similarly was removed because both failure and non-failure definitions were unique and also had suspended in definition.
4. Study ^80^ – removed because it also has “has results” in definition of success, which inflates the failure rate

**Merged**

*With Terminated, Withdrawn*

Studies ^38, 60^ (originally Terminated, Suspended). They fit well within the failure ranges of the studies.

*With Completed*

Study ^58^ (originally Completed AND the drug advanced to the next phase).

**New category**

*Completed, Ongoing/Active*

1. Any with 3/4 or 4/4 statuses listed under Ongoing:
   a. Studies ^14, 70^ (originally Completed, Active, "Good" Terminated),
   b. Study ^23^ (originally Completed, Active, not recruiting, Recruiting, Not yet recruiting),
   c. Studies ^49, 87^ (originally Completed, Ongoing).
2. Studies ^9, 16, 27, 28, 38, 44, 53, 54, 63, 77, 79^ (originally Completed, Ongoing, Unknown). We initially performed the analysis with these studies separately, however the analysis was not more informative, because the groups are too similar, which can be seen from fact that their distributions overlap:

|  | Median | Min | Max |
| --- | --- | --- | --- |
| Completed, Ongoing, Unknown | 10.75% | 4.30% | 15.97% |
| Completed, Ongoing/Active | 12.00% | 7.40% | 12.00% |

3. Study ^32^ (originally Completed, Active, Unknown). It is not explicit that Not Yet Recruiting was excluded, that status simply isn’t mentioned.

**eAppendix 4 Generalized linear model methodology**

**The choice of studies included in the model**

In order to analyze the impact of failure and non-failure definitions on failure proportions, the studies that were not analyzing Clinicaltrials.gov dataset were excluded. Also, studies with unique failure definitions and non-failure definitions among the studies analyzing Clinicaltrial.gov were excluded. Due to overlap with other therapeutic areas studies with no restriction on therapeutic area (including a paper analyzing AI across different disciplines, and a paper examining drug therapy trials in general) were excluded from the analysis. Similarly, due to overlap with many other studies, we excluded papers that general oncology, general gynecology, general surgery (including the paper analyzing robot-assisted surgery), and general pediatrics.

On the other hand, all studies without therapeutic area overlap with other studies were included in the model (topics: vascular surgery, cardiac surgery, dental implants, alcohol use disorder, connective tissue disease, obsessive compulsive disorder, posttraumatic stress disorder, ophthalmology, rare diseases, sepsis, COVID-19, pregnancy, pediatric obesity).

The remaining studies were divided into several groups based on therapeutic area overlap. When possible, the number of studies included from a group was maximized. When this was not plausible, the most representative study were chosen, based on the query date and sample size. The following groups were formed and the following studies chosen:

1. One group comprised two studies on urology, two studies on urooncology and a study on prostate cancer ^9, 47, 59, 70, 80^. Due to the fact that it was impossible to include any two studies from this group due to overlap, study ^9^ was chosen, as the studies on urologic oncology and prostate cancer ^47, 59, 70^ had a drastically lower number of included trials, and the other study on urology ^80^ had an older query of Clinicaltrials.gov
2. The second group included a study on head and neck cancer and a study on nasopharyngeal carcinoma ^22, 84^. Although the study on head and neck cancer concerned a broader topic ^84^ it utilized a much older query, so the study on nasopharyngeal carcinoma was included ^22^.
3. Two studies focused on gynecological oncology ^33, 76^, with study ^76^ being chosen since it had a more recent query.
4. Studies focusing on neurology-related topics included studies on neurology ^38^, deep brain stimulation ^62^, neurodegenerative diseases ^23^, Alzheimer's disease ^21^, and chronic pain ^83^. We included the studies ^21, 83^ since this was the only way to include more than one study due to overlap between other studies.
5. A study focused on heart failure ^19^ and another on cardiovascular trials ^65^, but the study on cardiovascular trials was an older query and had included less trials, so study ^19^ was included.
6. Seven studies analyzed orthopaedics and related topics. We excluded four studies due to overlap with other studies, in order to maximize the number of included studies: a study focusing on orthopaedic surgery ^71^, a study focusing on orthopaedic oncology ^18^, a study on upper and lower extremity fractures ^85^, and one on spinal diseases ^82^. Included studies were on hand and wrist ^10^, shoulder and elbow ^12^, spine ^13^. For the analysis, the shoulder and elbow portions of study ^12^ were separated, since stratified data was available for shoulder and for elbow trials and failure proportions. Therefore, this study was counted as two data points in our analysis.
7. Two studies focused on neuro-oncology ^8, 11^, one on neurosurgery ^48^, and one on glioblastoma ^68^. Since it was impossible to include more than one study from this group, the glioblastoma study was chosen ^68^ since it had the most recent query and more trials included than studies ^11, 40^ while study ^8^ included only US studies which could introduce bias.

**Variable categorization**

The variables included in the model were Failure Definition, which was categorized into four levels, with terminated, withdrawn, suspended set as reference; Non-failure Definition, categorized into three levels with completed as reference; Study Type, categorized into three levels with interventional set as reference; and Phases included, with any set as reference. The choice of reference categories was based on the highest observed frequency.

**Redundant success category handling**

Only two studies ^16, 81^ used the completed, active, not recruiting non-failure definition. Upon inspection of the manuscript of study ^81^, it was apparent that only 3 included trials (1.3%) had the active, not recruiting status. We excluded these 3 studies from the denominator of study ^81^ and reclassified it to completed as classification as a separate status is not reasonable for such a small proportion. Study ^16^ was excluded from the analysis since it now had a unique non-failure definition, and no stratified data was available on the number of studies with status completed and active, not recruiting.

**Model selection**

To address overdispersion identified in the initial binomial GLM (dispersion parameter = 13.93), a beta-binomial regression model with an overdispersion parameter (φ) was fitted using maximum likelihood estimation with the Broyden–Fletcher–Goldfarb–Shanno (BFGS) optimization algorithm to address convergence issues encountered with default methods. This model accounts for extra-binomial variance through φ, which quantifies residual heterogeneity not explained by the predictors. A sensitivity analysis was performed by removing non-significant predictors via backward selection.

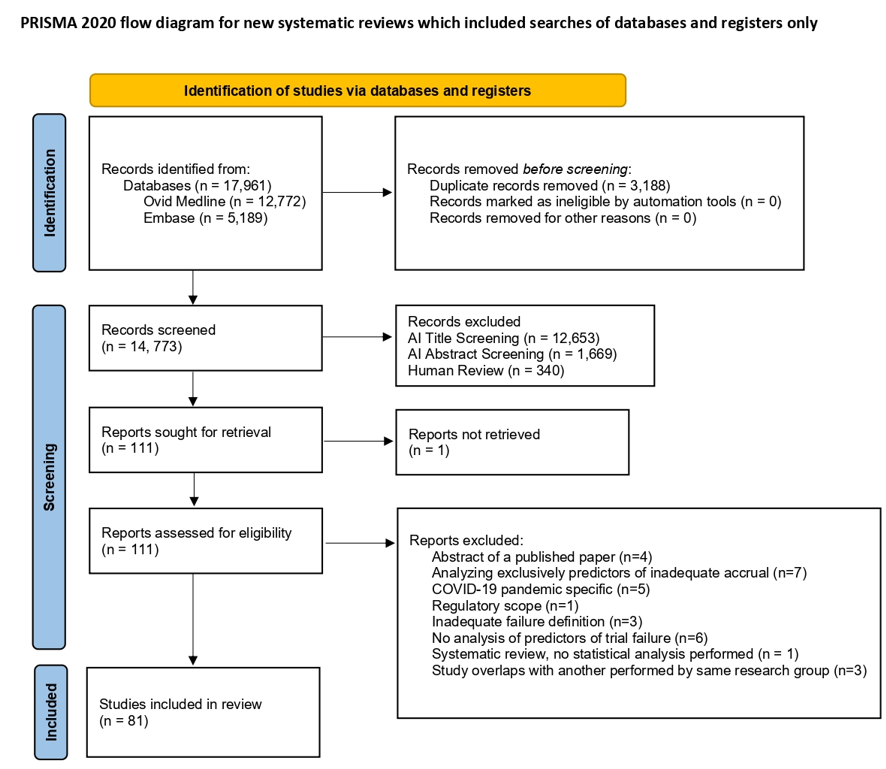

**eFigure 2 PRISMA Flowchart of the included studies**

**eTable 4 Characteristics of included studies**

| Study | Data source | Trials  (N) | Topic/ Demographics | Study type | Phase | Timeframe | Failed trials definition | Non-failed trials definition | Failed trials (%) |
| --- | --- | --- | --- | --- | --- | --- | --- | --- | --- |
| Smith et al. 2024 ^8^ | CT.gov | 1,257 | Neurooncology, adults^[1]^ | Any | N/A | Completed prior 31/12/2019 | Terminated, Withdrawn, Suspended | Completed | 26 |
| Magnani et al. 2021 ^9^ | CT.gov | 8,636 | Urology | Interventional | Any | Registered 01/10/2007-29/10/2019 | Terminated, Withdrawn, Suspended | Completed, Ongoing, Unknown | 11.60 |
| Delma et al. 2023 ^10^ | CT.gov | 793 | Hand and wrist | Any | N/A | Queried 03/05/2022 | Terminated | Completed | 9.70 |
| Butler et al. 2024 ^11^ | CT.gov | 139 | Neurooncology | RCT | 2/3, 3, 4 | Completed prior 01/03/2020 | Terminated, Withdrawn, Suspended, Unknown | Completed | 41 |
| Caruana et al. 2022a ^12^ | CT.gov | 788^[2]^ | Shoulder and elbow | Interventional | Any | Queried 06/08/2021 | Terminated | Completed | 8.60^[3]^ |
| Caruana et al. 2022b ^13^ | CT.gov | 969 | Spine | Interventional | Any | Queried 20/07/2021 | Terminated | Completed | 14 |
| Zhang et al. 2022 ^14^ | CT.gov | 567 | Gastric cancer | Interventional | 1/2, 2, 2/3, 3 | Started 01/01/2007-12/01/2020 | Terminated^[4]^ | Completed, Active, "Good" Terminated^[5]^ | 7.40^[6]^ |
| Lee et al. 2023 ^15^ | CT.gov | 209 | Oncology, older adults (≥60 years) | Interventional | 2-4 | Completed 2008-2019 | Terminated^[7]^, Withdrawn, Suspended, Completed with <85% of targeted sample size | Completed, Terminated^[8]^ | 41.60 |
| Luo et al. 2021 ^16^ | CT.gov | 298 | Obsessive-compulsive disorder | Interventional | Any | Registered prior 07/05/2020 | Terminated, Withdrawn, Suspended | Completed, Ongoing, Unknown | 9.40 |
| Weygandt et al. 2023 ^17^ | CT.gov | 54 | Posttraumatic stress disorder, military veterans^[9]^ | Interventional | Any | Started after 01/01/2000, completed prior 30/06/2018 | Terminated, Withdrawn, Suspended | Completed | 20.40 |
| Singh et al. 2024 ^18^ | CT.gov | 130 | Orthopaedic oncology | RCT | 3, 4 | Completed prior 01/05/2020 | Terminated, Withdrawn, Suspended, Unknown | Completed | 19.20 |
| Khan et al. 2021 ^19^ | CT.gov | 572 | Heart failure | RCT | Any | Completed prior 31/12/2017 | Terminated | Completed, Active, not recruiting | 21 |
| Yilmaz et al. 2019 ^20^ | CT.gov | 2,926 | Ophthalmology | Interventional | Any | Queried 30/05/2019 | Terminated, Withdrawn, Suspended | Completed | 14 |
| Yilmaz et al. 2018 ^21^ | CT.gov | 744 | Alzheimer's disease and mild cognitive impairment | Interventional | Any | Queried 15/01/2018 | Terminated, Withdrawn, Suspended | Completed | 17 |
| Huang et al. 2024 ^22^ | CT.gov | 311 | Nasopharyngeal carcinoma | Interventional | Any | Queried 01/01/2023 | Terminated, Withdrawn, Suspended | Completed | 18.00 |
| Stefaniak et al. 2017 ^23^ | CT.gov | 362 | Neurodegenerative diseases, adults^[10]^ | RCT | 2, 2/3, 3, 4 | Registered 01/01/2000-31/12/2009 | Terminated, Withdrawn, Suspended | Completed, Active, not recruiting, Recruiting, Not yet recruiting | 12 |
| Amstutz et al. 2017 ^24^ | SNSF project database | 101 | General | RCT | Any | Recruitment and funding ended by 30/04/2015 | Discontinued^[11]^ | Completed | 26 |
| Pica et al. 2016 ^25^ | CT.gov | 559 | Pediatrics | RCT | Any | Registered 01/01/2008-31/12/2010, completed prior 31/12/2012 | Terminated, Withdrawn, Suspended | Completed | 19 |
| Chapman et al. 2014 ^26^ | CT.gov | 395 | Surgery, adults | RCT | 3, 4 | Registered 01/01/2008-31/12/2009, final confirmed status prior 31/12/2011 | Terminated, Withdrawn, Suspended | Completed | 21 |
| Mouw et al. 2018 ^27^ | CT.gov | 88,498^[12]^ | General^[13]^ | Any | N/A | Registered 2005-2015 | Terminated, Withdrawn | Completed, Ongoing, Unknown | 15.97^[14]^ |
| Long et al. 2023 ^28^ | CT.gov | 529 | Robot-assisted surgery^[15]^ | Interventional | Any | Queried 25/05/2021 | Terminated, Withdrawn, Suspended | Completed, Ongoing, Unknown | 10.20 |
| Brewster et al. 2022 ^29^ | CT.gov | 13,259 | Pediatrics | Interventional | Any | Registered 01/10/2007-09/03/2020 | Terminated, Withdrawn, Suspended | Completed | 11.10 |
| Zhang et al. 2023 ^30^ | CT.gov | 8,687 | Oncology^a^ | Interventional | Any | Registered 27/09/2007-30/06/2015 | Terminated | Completed, Active, not recruiting, Recruiting, Enrolling by invitation, Suspended | 22.74 |
| Brigante et al. 2023 ^31^ | CT.gov | 175 | Connective tissue diseases | RCT | 2/3, 3, or 4 | Registered prior 19/03/2021, started after 01/01/2000 | Terminated, Withdrawn, Suspended, Unknown | Completed | 33 |
| Aziz et al. 2024 ^32^ | MEDLINE | 98 | Hyperacute stroke^[16]^ | RCT | 2, 3 | Published in 9 prespecified major clinical journals 01/01/2013-31/12/2022 | Prematurely terminated^[17]^ | Nonterminated | 46 |
| Swailes et al. 2019 ^33^ | CT.gov | 318 | Gynecological oncology | RCT | Any | Registered 01/01/2009-31/12/2013 | Terminated, Withdrawn, Suspended | Completed, Active, Unknown | 12 |
| Nguyen et al. 2018 ^34^ | CT.gov | 134 | Radiation oncology^[18]^ | RCT | Any | Registered 27/09/2007-31/12/2012 | Terminated, Withdrawn | Completed | 29.90 |
| Kavalci et al. 2023 ^35^ | CT.gov | 112,647 | General | Interventional | Any | Registered Jan 2011-July 2022 | Terminated, Withdrawn | Completed | N/A^[19]^ |
| Chao et al. 2021 ^36^ | CT.gov | 284,644 | General^[20]^ | Any | N/A | Queried 02/10/2018 | Terminated | Completed, Ongoing, Unknown, Suspended, Withdrawn | 5.60 |
| Geletta et al. 2019 ^37^ | CT.gov | 119,591 | General | Any | N/A | Started prior 01/05/2015 | Terminated | Completed | 9.99 |
| Turner et al. 2021 ^38^ | CT.gov | 16,994 | Neurology | Interventional | Any | Registered 01/10/2007-30/04/2018 | Terminated, Suspended | Completed, Ongoing, Unknown | 11.30 |
| Rees et al. 2019 ^39^ | CT.gov | 659 | Rare diseases^[21]^ | RCT | Any | Registered 01/01/2010-31/12/2012, completed prior 31/12/2014 | Terminated, Withdrawn, Suspended | Completed | 30.20 |
| Speich et al. 2022 ^40^ | REC Protocols | 326 | General | RCT | 2-4 | RCT protocols approved in 2012 by RECs located in Switzerland, the UK, Germany, and Canada | Discontinued^[22]^ | Completed, Unclear^[23]^ | 30 |
| Kim et al. 2022 ^41^ | CT.gov | 819 | Pregnancy^[24]^ | Interventional | Any | Started 01/01/2009-31/12/2018 | Terminated, Withdrawn | Completed, Ongoing, Unknown, Suspended | 12.90 |
| Elkin et al. 2021a ^42^ | CT.gov | 68,999 | General^[25]^ | Any | N/A | Started after 01/01/2000, queried May 2019 | Terminated | Completed | 11.46 |
| Cancelli et al. 2024 ^43^ | CT.gov | 746 | Cardiac surgery | Interventional | Any | Registered 01/01/1991-31/12/2023 | Terminated, Withdrawn, Suspended | Completed | 22.70 |
| Khunger et al. 2018 ^44^ | CT.gov | 12,875 | Oncology, adults | RCT | Any | Registered 16/11/2011-16/04/2015 | Terminated, Withdrawn | Completed, Ongoing, Unknown | 8.50 |
| Filippi-Arriaga et al. 2023 ^45^ | REec | 3,419 | General | Interventional | Any | Registered 01/01/2013-31/11/2021 | Prematurely terminated | Terminated as planned | 21 |
| Hedenmalm et al. 2011 ^46^ | EudraCT, LVIS-C, and Documentum | 772^[26]^ | General | Interventional | 3 | Prematurely ended 01/01/2002-31/12/2008 | Prematurely terminated^[27]^ | All other CT applications | 8^[28]^ |
| Stensland et al. 2021 ^47^ | CT.gov | 293 | Prostate cancer^[29]^ | Interventional | 2, 3 | Registered Jan 2007-Dec 2020 | Terminated | Completed | 32.80 |
| Jamjoom et al. 2017 ^48^ | CT.gov | 64 | Neurosurgery^[30]^ | RCT | 3, 4 | Registered 01/01/2000-31/12/2012 | Terminated, Withdrawn, Suspended | Completed | 26.60 |
| Dufetelle et al. 2018 ^49^ | CT.gov | 65,095 | General | RCT | Any | Registered 29/02/2000-31/12/2016 | Terminated, Withdrawn, Suspended | Completed, Ongoing | 10.00^[31]^ |
| Hartwell et al. 2022 ^50^ | CT.gov | 87 | Alcohol use disorder, Drug clinical trials^cc^ | Interventional | Any | Completed 01/10/2008-30/09/2018 | Terminated, Withdrawn | Completed | 12.60 |
| Shieh et al. 2022 ^51^ | CT.gov | 3,623 | Pregnancy | Any | N/A | Queried 13/10/2020 | Terminated | Completed | 9.20^[32]^ |
| van den Bogert et al. 2017 ^52^ | ToetsingOnline database^[33]^ | 574 | Drug clinical trials | Interventional | Any | Approved by Dutch IRBs in 2007 | Discontinued | Completed as planned | 17.80 |
| Hu et al. 2023 ^53^ | CT.gov | 1,061 | Sepsis | Interventional | Any | Registered prior 08/07/2022 | Terminated, Withdrawn, Suspended | Completed, Ongoing, Unknown | 12.00 |
| Wang et al. 2022 ^54^ | CT.gov | 1,312 | Obesity, Pediatrics | Interventional | Any | Registered prior 29/07/2021 | Terminated, Withdrawn, Suspended | Completed, Ongoing, Unknown | 4.30 |
| Binko et al. 2023 ^55^ | CT.gov | 108 | Vascular surgery, adults | RCT | 2/3, 3, 4 | Registered 01/01/2010-31/10/2019, completed prior to 31/10/2019 | Terminated, Withdrawn, Suspended | Completed | 22.20 |
| Chen et al. 2024 ^56^ | CT.gov | 536 | Artificial intelligence^[34]^ | Interventional | Any | Queried 23/12/2023 | Terminated, Withdrawn, Suspended | Completed | 9.50 |
| Elkin et al. 2021b ^57^ | CT.gov | 772 | COVID-19 | Interventional | Any | Queried Jan 2021 | Terminated, Withdrawn, Suspended | Completed | 18.65 |
| Chang et al. 2023 ^58^ | CT.gov | 18,304 | Oncology, Drug clinical trials | Interventional | 1-3 | Queried 24/07/2022 | Terminated, Withdrawn, Suspended | Completed AND the drug advanced to the next phase | 38.10 |
| Alhajahjeh et al. 2024 ^59^ | CT.gov | 1,033 | Urologic oncology | Interventional | Any | Results posted prior Dec 2020 | Terminated, Withdrawn, Suspended | Completed, Active, not recruiting | 25.38 |
| Buergy et al. 2020 ^60^ | CT.gov | 1,482 | Oncology | Interventional | 3 | Registered after Jan 2006, updated prior May 2017 | Terminated, Suspended^[35]^ | Completed | 23.30 |
| Lee et al. 2020 ^61^ | CT.gov | 317 | Dental implants | Any | N/A | Queried 29/05/2020 | Terminated, Withdrawn, Suspended | Completed | 9.50 |
| Mishra et al. 2024 ^62^ | CT.gov | 325 | Deep brain stimulation | Any | N/A | Queried Dec 2022 | Terminated, Withdrawn | Completed | 20.30 |
| Bao et al. 2023 ^63^ | CT.gov | 225 | Contrast-enhanced ultrasound ^[36]^ | Any | N/A | Queried 22/04/2022 | Terminated, Withdrawn, Suspended | Completed, Ongoing, Unknown | 14.22 |
| Luo et al. 2023 ^64^ | CT.gov | 12,717 | Therapy-related | Any | N/A | Not listed | Terminated, Withdrawn | Completed | 44.80^[37]^ |
| Roddick et al. 2017 ^65^ | CT.gov | 431 | Coronary artery disease, acute coronary syndrome, heart failure and atrial fibrillation | Interventional | 1/2, 2, 2/3, 3, 4 | Completed 01/01/2010-01/01/2014 | Terminated, Withdrawn, Suspended | Completed | 17.90 |
| Huo et al. 2021 ^66^ | DTRIPP | 312 | Drug clinical trials | Interventional | Any | Registered prior 31/03/2020 | Terminated^[38]^, Stopped^[39]^ | Completed, Ongoing | 3.00 |
| Tanemura et al. 2021 ^67^ | EudraCT | 142 | Pediatrics^[40]^ | Interventional | 2, 2/3, 3 | Registered 01/01/2014-31/12/2018 | Not authorized^[41]^, Temporarily halted^[42]^, Prematurely ended^[43]^ | All cases completed | 33.10 |
| Shah et al. 2022 ^68^ | CT.gov | 886 | Glioblastoma | Interventional | Any | Queried 31/03/2022 | Terminated | Completed | 19.80 |
| Abdulelah et al. 2024 ^69^ | CT.gov | 240 | Pulmonary hypertension | Interventional | Any | Started after 01/01/2020, completed prior 31/12/2020 | Terminated | Completed, Active, not recruiting | 30.40 |
| Stensland et al. 2021 ^70^ | CT.gov | 1,869 | Urologic oncology, adults | Interventional | 1/2, 2, 2/3, 3 | Started 01/01/2007-20/04/2019 | Terminate^d^ | Completed, Active, "Good" Terminated^[44]^ | 12.00^[45]^ |
| Hecht et al. 2023 ^71^ | CT.gov | 8,603 | Orthopaedic surgery | Interventional | Any | Started 02/10/2007-07/10/2022 | Terminated, Withdrawn, Suspended | Completed | 16 |
| Steinberg et al. 2024 ^72^ | CT.gov | 3,087 | Obstetrics and gynecology^a^ | Interventional | Any | Registered 01/10/2007-09/03/2020 | Terminated, Withdrawn, Suspended | Completed, Ongoing, Unknown | 9.30 |
| Steinberg et al. 2022 ^73^ | CT.gov | 8,174 | Gynecology | Interventional | Any | Registered after 01/10/2007, completed prior 08/03/2017 | Terminated, Withdrawn, Suspended | Completed, Ongoing, Unknown | 9.50 |
| Demetriades et al. 2022 ^74^ | CT.gov | 112 | Spine | RCT | 3-4 | Screened 01/01/2013-31/12/2020 | Terminated, Withdrawn, Suspended | Completed | 22 |
| Jacobsen et al. 2023 ^75^ | CT.gov | 408 | Chronic pain | RCT | 2/3, 3, 4 | Completed prior 01/10/2017 | Terminated, Withdrawn, Suspended, Unknown | Completed | 27.50 |
| Johnson et al. 2019 ^76^ | CT.gov | 130 | Head and neck cancer | RCT | 2/3, 3, 4 | Completed prior 01/03/2016 | Terminated, Withdrawn, Suspended, Unknown | Completed | 29.20 |
| Shepard et al. 2023 ^77^ | CT.gov | 142 | Extremity fractures | RCT | 2/3, 3, 4 | Completed prior 09/09/2017 | Terminated, Withdrawn, Suspended, Unknown | Completed | 40.10 |
| Steele et al. 2020 ^78^ | CT.gov | 653 | Atopic eczema | Any | N/A | Queried 23/05/2019 | Terminated, Withdrawn, Unknown | Completed, Ongoing, Suspended | 13.80 |
| Stensland et al. 2014 ^79^ | CT.gov | 7,776 | Oncology, adults | Interventional | 1/2, 2, 2/3, 3 | Registered 11/9/2005-11/11/2011 | Terminated, Withdrawn | Completed, Ongoing | 12 |
| Bandari et al. 2020 ^80^ | CT.gov | 1,340 | Urology | Interventional | Any | Screened 01/01/2006-31/12/2015 | Terminated, Withdrawn | Completed, Has Results | 46.10 |
| Lu et al. 2024 ^81^ | CT.gov | 85 | Cervical cancer and precancer | RCT | 3-4 | Started after 01/01/2020, completed prior 31/12/2020 | Terminated, Withdrawn, Suspended, Unknown, Completed^[46]^ | Completed, Active, not recruiting, Unknown | 51.80% |
| Yadete et al. 2024 ^82,[47]^ | CT.gov | 1,784 | Inflammatory bowel disease | Any | N/A | Listed Jan 1996-Jun 2023 | Failed^[48]^ | Completed | 23% |
| Togra et al. 2023 ^83^,^uu^ | CT.gov | 4,272 | Heart failure | Any | N/A | Registered 27/9/2007-31/3/2022 | Terminated^[49]^, Withdrawn, Suspended | Not stated^[50]^ | 11.52% |
| Aljabali et al. 2023 ^84^,^uu^ | CT.gov | 3,946 | Cardiovascular | Interventional | Any | Completed Jan 2000-June 2022 | Discontinued before completion^vv^ | Not stated^[51]^ | 18.50% |
| Aljabali et al. 2023 ^85^,^uu^ | CT.gov | 1,181 | Heart failure | Interventional | Any | Completed prior June 2022 | Discontinued before completion^vv^ | Not stated^[52]^ | 24.50% |
| Spinosa et al. 2021 ^86^,^uu^ | CT.gov | 1,882 | Gynecological oncology | Interventional | Any | Queried Feb 2020 | Terminated, Withdrawn, Suspended | Completed | 21% |
| Shahait et al. 2024 ^87^,^uu^ | CT.gov | 202 | Nephrolithiasis | Any | N/A | Completed prior 31/12/2020 | Terminated^[53]^ | Completed^[54]^ | 17.30% |
| Rotter et al. 2025 ^88, [55]^ | CT.gov | 3,522 | Leukemia | Interventional | Any | 2000-2020 | Prematurely terminated^vv^ | Not stated | 28.40% |

Abbreviations: CT.gov – Clinicaltrials.gov; SNSF - Swiss National Science Foundation; RCT - Randomized Controlled Trial; RECs - Research Ethics Committees; REec - Spanish Registry of Clinical Studies; IRB - Institutional review board; DTRIPP - Drug Trial Registration and Information Publication Platform (China); EudraCT - European Clinical Trials Database.

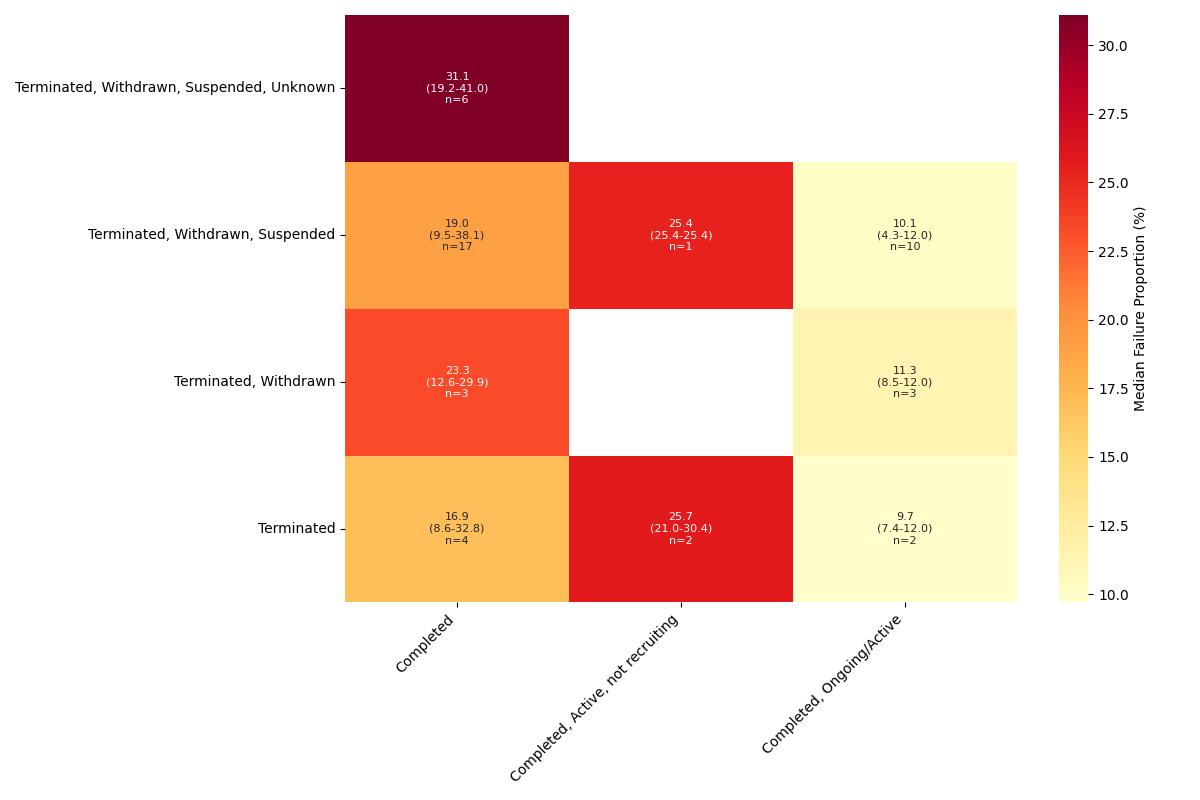

**eFigure 3 Failure proportion distribution by trial failure and non-failure definitions, studies including interventional studies only, median (range)**

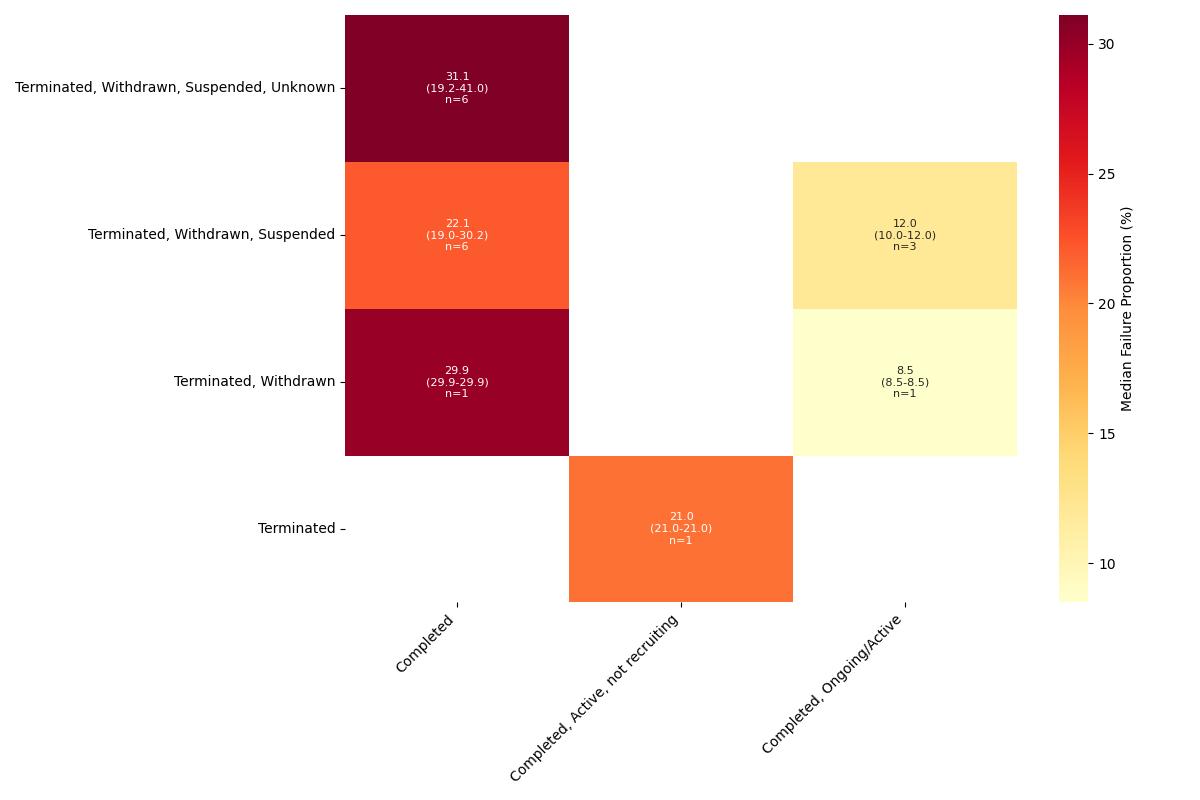

**eFigure 4 Failure proportion distribution by trial failure and non-failure definitions, studies including randomized controlled trials only, median (range)**

**eTable 5 Statistically significant predictors of study failure in the multivariable analysis**

| Study | Multivariable analysis | Sample size | Location | Number of centers | Blinding/  Randomization | Phase | Funding source | Primary purpose | Intervention type | Other | Covariates in multivariable analysis |
| --- | --- | --- | --- | --- | --- | --- | --- | --- | --- | --- | --- |
| Magnani et al. 2021 ^9^ | Cox regression | 0-9 vs 100-499: (HR=58.83, 95% CI 44.13-78.41, p<0.001)  10-49 vs 100-499: (HR=6.92, 95% CI 5.17-9.26, p<0.001)  50-99 vs 100-499: (HR=1.94, 95% CI 1.36-2.78, p<0.001) | None significant | Multicenter vs single center: (HR=1.18, 95% CI 1.01-1.39, p=0.038) | None significant/  None significant | Phase 1 vs Phase 2/3-3: (HR=0.48, 95% CI 0.36-0.65, p<0.001)  Not Applicable vs Phase 2/3-3: (HR=0.74, 95% CI 0.56-0.97, p=0.029) | Academic vs Industry: (HR=0.81, 95% CI 0.69-0.96, p=0.012)  Government vs Industry: (HR=0.62, 95% CI 0.49-0.78, p<0.001) | Diagnostic vs Treatment: (HR=1.49, 95% CI 1.17-1.91, p=0.001) | Procedure vs Pharmaceutical: (HR=0.69, 95% CI 0.52-0.92, p=0.012)  Other vs Pharmaceutical: (HR=0.66, 95% CI 0.50-0.87, p=0.003) | Endourology vs Oncology: (HR=2.00, 95% CI 1.35-2.95, p<0.001)  General vs Oncology: (HR=1.24, 95% CI 0.99-1.56, p=0.061)  First Submission Year: (HR=1.02, 95% CI 1.00-1.05, p=0.044) | Sample size, location, number of centers, blinding, randomization, phase, funding source, primary purpose, intervention type, number of arms, first submission year, data monitoring committee |
| Butler et al. 2024 ^11^ | Logistic regression | Didn't investigate | None significant | Didn't investigate | Didn't investigate/ Didn’t investigate | Didn't investigate | Other funding vs Industry/NIH: (OR=4.24, 95% CI 1.31-13.78, p<0.05) | Didn't investigate | Device vs Biological: (OR=0.04, 95% CI 0.00-0.83, p<0.05)  Procedure vs Biological:  (OR=0.02, 95% CI 0.00-0.47, p<0.05)  Drug+procedure vs Biological:  (OR=0.02, 95% CI 0.00-0.60, p<0.05)  Drug+radiation vs Biological:  (OR=0.07, 95% CI 0.01-0.93, p<0.05)  Multiple interventions vs Biological:  (OR=0.03, 95% CI 0.00-0.59, p<0.05) | Didn't investigate | Location, funding source, intervention type |
| Caruana et al. 2022a ^12^ | Logistic regression | Didn't investigate | Didn't investigate | Didn't investigate | Blinded vs not blinded: (OR=45.8, 95% CI 9.1-841.4, p=0.0003)/  None significant | None significant | Industry vs local groups sponsorship: (OR=4.2, 95% CI 1.7-10.0, p=0.001) | Didn't investigate | None significant | Didn't investigate | Blinding, randomization, phase, funding source, intervention type |
| Caruana et al. 2022b ^13^ | Logistic regression | Didn't investigate | Didn't investigate | Didn't investigate | None significant/  None significant | Phase II vs Phase III: (OR=2.845, 95% CI 1.479-5.537, p=0.002) | Industry vs local groups: (OR=1.61, 95% CI 1.043-2.496, p=0.032) | Didn't investigate | Device vs drug/biological: (OR=2.213, 95% CI 1.226-3.971, p=0.008) |  | Blinding, randomization, phase, funding source, intervention type |
| Zhang et al. 2022 ^14^ | Logistic regression | Interaction with number of centers investigated | Didn't investigate | Interaction with anticipated accrual investigated | Didn't investigate/  Didn't investigate | Not significant in univariate analysis, excluded from multivariate | Not significant in univariate analysis, excluded from multivariate | Didn't investigate | Not significant in univariate analysis, excluded from multivariate | Anticipated accrual in single centre trials: (OR=0.60, 95% CI 0.41-0.88, p=0.009)  Anticipated accrual in multi-centre trials: (OR=0.72, p=0.025) | Interaction: Anticipated accrual x number of centers |
| Lee et al. 2023 ^15^ | Logistic regression | None significant | Non-US only vs US only: (OR=0.32, 95% CI 0.12-0.82, p<0.05) | Per 10 additional centers: (OR=0.83 , 95% CI 0.71-0.94, p<0.05) | Didn't investigate/  Didn't investigate | Didn't investigate | None significant | Didn't investigate | Didn't investigate | Life expectancy restriction vs no restriction: (OR=2.17, 95% CI 1.04-4.73, p<0.05) | Location, number of centers, blinding, randomization, phase, funding source, target sample size, number of countries, number of arms, first-line setting, disease-related factors, endpoint-related factors, intervention-related factors, eligibility-related factors |
| Luo et al. 2021 ^16^ | Cox regression | Not significant in univariate analysis, excluded from multivariate | Didn't investigate | Didn't investigate | Not significant in univariate analysis, excluded from multivariate/  Not significant in univariate analysis, excluded from multivariate | Not significant in univariate analysis, excluded from multivariate | Significant in univariate analysis, collinearity found with variable "interventions" so excluded from multivariate analysis | Not significant in univariate analysis, excluded from multivariate | Drug trials vs non-drug trials: (HR=3.93, 95% CI: 1.71-9.08, p=0.001) | Absence of collaborators vs presence: (HR=5.17, 95% CI: 1.97-13.54, p=0.001) | Intervention type, collaborators |
| Singh et al. 2024 ^18^ | Logistic regression | ≥261 vs <261: (OR=0.85, 95% CI 0.42-0.95, p<0.05) | Didn't investigate | Didn't investigate | Didn't investigate/  Didn't investigate | Didn't investigate | None significant | Didn't investigate | None significant | Didn't investigate | Sample size, funding source, intervention type |
| Khan et al. 2021 ^19^ | Logistic regression | Didn't investigate | Didn't investigate | Didn't investigate | Completion: Triple blind vs open label: (OR=1.88, 95% CI 1.01-3.50, p=0.04)/  Didn't investigate | None significant | None significant | Didn't investigate | Completion: Behavioural vs drug intervention: (OR=0.24, 95% CI 0.07-0.79, p=0.02)  Device vs drug intervention: (OR=1.72, 95% CI 1.10-2.70, p=0.02 | Didn't investigate | Blinding, phase, funding source, intervention type |
| Huang et al. 2024 ^22^ | Logistic regression | None significant | Didn't investigate | Didn't investigate | Didn't investigate/  None significant | None significant | Industry vs Nonindustry funding: (OR=3.12, 95% CI 1.42-6.82, p=0.003) | Didn't investigate | None significant | Recurrent/metastatic vs Early/intermediate setting: (OR=11.95, 95% CI 2.27-62.93, p=0.003) | Sample size, randomization, phase, funding source, intervention type, disease setting |
| Stefaniak et al. 2017 ^23^ | Logistic regression | ≤100 vs >100: (OR=2.65, 95% CI 1.21-5.78, p=0.015) | Didn't investigate | Not significant in univariate analysis, excluded from multivariate | Not significant in univariate analysis, excluded from multivariate/  Didn't investigate | Phase IV vs Phase III trials: (OR=3.90, 95% CI 1.41-10.83, p=0.009) | None significant | Didn't investigate | None significant | Didn't investigate | Sample size, blinding, phase |
| Amstutz et al. 2017 ^24^ | Logistic regression | None significant | Didn't investigate | None significant | Didn't investigate/  Didn't investigate | Didn't investigate | Didn't investigate | Didn't investigate | Didn't investigate | Didn't investigate | Number of centers, target sample size, start year |
| Pica et al. 2016 ^25^ | Logistic regression | Planned sample size: (OR=0.999, 95% CI 0.998-1.00, p=0.008) | Didn't investigate | Didn't investigate | None significant/  Didn't investigate | Didn't investigate | Academic vs Industry funding: (OR=0.46, 95% CI 0.27-0.77, p=0.004) | Didn't investigate | None significant | Didn't investigate | Blinding, funding source, intervention type, target sample size |
| Chapman et al. 2014 ^26^ | Logistic regression | None significant | Didn't investigate | Didn't investigate | Didn't investigate/  Didn't investigate | Didn't investigate | None significant | Didn't investigate | None significant | Didn't investigate | Sample size, funding source, intervention type |
| Long et al. 2023 ^28^ | Cox regression | Didn't investigate | Didn't investigate | Didn't investigate | None significant/  Didn't investigate | Didn't investigate | NIH vs Industry funding: (HR=3.30, 95% CI 1.09-10.00, p=0.04) | Device feasibility vs Treatment: (HR=2.30, 95% CI 1.08-4.89, p=0.03) | Didn't investigate | Didn't investigate | Blinding, funding source, primary purpose, allocation model |
| Brewster et al. 2022 ^29^ | Logistic regression | 10-49 vs 100-499: (OR=4.62, 95% CI 3.47-6.18)  50-99 vs 100-499: (OR=1.90, 95% CI 1.38-2.61) | High-income country only vs Low- to middle-income country only: (OR=1.87, 95% CI 1.47-2.66)  Low- to middle-income country and high-income country vs Low- to middle-income country only: (OR=2.10, 95% CI 1.27-3.47) | None significant | None significant/  Randomized vs Not: (OR=2.53, 95% CI 1.94-3.31) | Phase 1 vs 2/3-3: (OR=0.44, 95% CI 0.30-0.66) | Government vs industry funding: (OR=0.72, 95% CI 0.47-0.97)  Academic vs industry funding: (OR=0.64, 95% CI 0.50-0.82)  Other vs industry funding: (OR=0.63, 95% CI 0.41-0.96) | Didn't investigate | Behavioral vs Not: (OR=0.64, 95% CI 0.42-0.98)  Drugs, biologics, supplements vs Not: (OR=1.51, 95% CI 1.05-2.17) | Data monitoring committee present vs Not: (OR=1.29, 95% CI 1.06-1.56) | Sample size, number of centers, blinding, randomization, phase, funding source, intervention type, data monitoring committee, therapeutic focus, location (income) |
| Zhang et al. 2023 ^30^ | Logistic regression | Didn't investigate | US and international vs US only: (OR=0.65, 95% CI 0.55-0.76, p<0.001) | Didn't investigate | Didn't investigate/  Not significant in univariate analysis, excluded from multivariate | Phase 2 vs Phase 1: (OR=1.27, 95% CI 1.14-1.41, p<0.001) | Other funding vs Industry funding: (OR=1.19, 95% CI 1.02-1.38, p=0.025) | Didn't investigate | Not significant in univariate analysis, excluded from multivariate | Multiple histology vs Hematologic cancer: (OR=0.70, 95% CI 0.54-0.91, p=0.007) | Location, phase, funding source, cancer type |
| Brigante et al. 2023 ^31^ | Logistic regression | None significant | Didn't investigate | Didn't investigate | Didn't investigate/  Didn't investigate | None significant | Didn't investigate | Didn't investigate | None significant | Didn't investigate | Phase, intervention type, target sample size, disease |
| Aziz et al. 2024 ^32^ | Logistic regression | Didn't investigate | Not significant in univariate analysis, excluded from multivariate | Not significant in univariate analysis, excluded from multivariate | Not significant in univariate analysis, excluded from multivariate/  Didn't investigate | Not significant in univariate analysis, excluded from multivariate | Not significant in univariate analysis, excluded from multivariate | Didn't investigate | Didn't investigate | Complex trial vs non-complex: (aOR=2.76, 95% CI 1.13-7.49)  Presence of futility rule vs absence: (aOR=4.43, 95% CI 1.62-17.91)  None/slight prestroke disability only vs dependency allowed: (aOR=2.19, 95% CI 0.84-6.72) | Trial complexity, futility rule, prestroke dependency |
| Nguyen et al. 2018 ^34^ | Logistic regression | Didn't investigate | Not significant in univariate analysis, excluded from multivariate | Not significant in univariate analysis, excluded from multivariate | None significant/  Didn't investigate | None significant | Not significant in univariate analysis, excluded from multivariate | Didn't investigate | Surgery vs non-surgical: (OR=12.30, 95% CI 1.38-109.9, p=0.025)  Behavioral vs non-behavioral: (OR=0.11, 95% CI 0.01-0.95, p=0.045) | Not reported vs efficacy endpoint (OR=4.44, 95% CI 1.37-14.35, p=0.013) | Intervention type, endpoint type |
| Geletta et al. 2019 ^37^ | Logistic regression | 0-100 vs >1000: (OR=3.96, 95% CI 3.47-4.54, p<0.05)  101-1000 vs >1000: (OR=1.56, 95% CI 1.36-1.80, p<0.05) | Didn't investigate | Didn't investigate | Didn't investigate/  Didn't investigate | Didn't investigate | Didn't investigate | Didn't investigate | Didn't investigate | Surgery topic: (OR=1.90, 95% CI 1.74-2.07, p<0.05)  Dermatological topic: (OR=0.08, 95% CI 0.06-0.10, p<0.05)  Coronary topic: (OR=0.15, 95% CI 0.11-0.20, p<0.05)  HIV/Pregnancy topic: (OR=0.21, 95% CI 0.18-0.24, p<0.05) | Sample size, topic probabilities |
| Turner et al. 2021 ^38^ | Cox regression | 0-9 vs 100-499: (HR=26.65, 95% CI 22.33-31.80, p<0.0001)  10–49 vs 100-499: (HR=2.74, 95% CI 2.29-3.27, p<0.0001)  50–99 vs 100-499: (HR=1.35, 95% CI 1.08-1.68, p=0.008) | Only high-income countries vs mixed/low-income countries: (HR=1.42, 95% CI 1.09-1.86, p=0.0096) | Multicenter vs single center: (HR=1.22, 95% CI 1.08-1.38, p=0.001) | Double vs no blinding: (HR=1.26, 95% CI 1.07-1.49, p=0.005)/  Randomized vs non-randomized trials: (HR=1.48, 95% CI 1.27-1.72, p<0.0001) | Phase 1 vs Phase 2/3–3: (HR=0.56, 95% CI 0.47-0.68, p<0.0001)  Phase 1/2–2 vs Phase 2/3–3: (HR=0.76, 95% CI 0.65-0.88, p=0.0003)  Phase 4 vs Phase 2/3–3: (HR=0.65, 95% CI 0.54-0.80, p<0.0001) | Academic vs industry funding: (HR=0.56, 95% CI 0.50-0.63, p<0.0001)  US Government vs industry funding: (HR=0.46, 95% CI 0.39-0.55, p<0.0001) | Other vs Treatment (HR=1.21, 95% CI 1.05–1.40) | Drugs/supplements vs other interventions: (HR=1.33, 95% CI 1.12-1.58, p=0.001)  Procedure vs other interventions: (HR=1.28, 95% CI 1.07–1.53, p=0.006)  Device vs other interventions: (HR=2.82, 95% CI 1.97-4.03, p<0.0001 | Neurodegenerative vs other conditions: (HR=1.40, 95% CI 1.17-1.67, p=0.0003)  Neurovascular vs other conditions: (HR=1.37, 95% CI 1.18-1.59, p<0.0001)  Pain vs other conditions: (HR=1.66, 95% CI 1.36-2.02, p<0.0001)  Inflammation vs other conditions: (HR=1.23, 95% CI 1.03-1.48, p=0.026)  Data monitoring committee present vs Not: (HR=0.84, 95% CI 0.76-0.93, p=0.001) | Sample size, number of centers, blinding, randomization, phase, funding source, primary purpose, intervention type, first submission year, data monitoring committee, disease, location (income) |
| Rees et al. 2019 ^39^ | Logistic regression | Didn't investigate | Didn't investigate | Didn't investigate | None significant/  Didn't investigate | None significant | Healthcare center vs industry funding: (OR=2.42, 95% CI 1.34-4.39, p=0.003)  Other funding vs industry funding: (OR=2.79, 95% CI 1.33-5.86, p=0.007) | Didn't investigate | None significant | Didn't investigate | Blinding, phase, intervention type, age group |
| Speich et al. 2022 ^40^ | Logistic regression | None significant | Didn't investigate | None significant | Didn't investigate/  Didn't investigate | Didn't investigate | None significant | Didn't investigate | Didn't investigate | Didn't investigate | Number of centers, industry sponsorship, target sample size, placebo control, reported recruitment projection, proportion of SPIRIT items reported in the protocol |
| Kim et al. 2022 ^41^ | Cox regression | Completion: Planned sample size n≥329 vs 0 ≤n< 80: (HR=0.53, 99% CI 0.36-0.77, p<0.01) | Completion: Very high HDI country vs Low HDI country: (HR=0.28, 99% CI 0.15-0.49, p<0.01) | Didn't investigate | Not significant in univariate analysis, excluded from multivariate/  Not significant in univariate analysis, excluded from multivariate | Not significant in univariate analysis, excluded from multivariate | Not significant in univariate analysis, excluded from multivariate | Didn't investigate | Didn't investigate | Completion: Abortion vs other: (HR=3.30, 99% CI 1.92-5.69, p<0.01)  Labor vs other: (HR=2.16, 99% CI 1.55-3.03, p<0.01)  Iron deficiency anemia vs other: (HR=2.29, 99% CI 1.44-5.92, p<0.01) | Target sample size, disease, location (income) |
| Cancelli et al. 2024 ^43^ | Logistic regression | None significant | Asia vs North America: (OR=0.18, 95% CI 0.07-0.39, p<0.001)  Europe vs North America: (OR=0.49, 95% CI 0.30-0.80, p=0.004)  Others vs North America: (OR=0.16, 95% CI 0.02-0.58, p=0.017) | Didn't investigate | Didn't investigate/  Didn't investigate | Phase 2 vs Phase 1: (OR=3.02, 95% CI 1.31-7.93, p=0.015)  Phase 4 vs Phase 1: (OR=3.62, 95% CI 1.43-10.23, p=0.010) | None significant | Didn't investigate | None significant | LVAD vs CABG: (OR=3.65, 95% CI 1.65-8.00, p=0.001)  Valve surgery vs CABG: (OR=4.30, 95% CI 2.33-8.00, p<0.001)  Aortic surgery vs CABG: (OR=2.86, 95% CI 1.22-6.43, p=0.012) | Sample size, location, phase, funding source, intervention type, year of enrollment, surgical procedure |
| Khunger et al. 2018 ^44^ | Cox regression | Didn't investigate | Didn't investigate | Didn't investigate | Didn't investigate/  Didn't investigate | Phase II vs Phase I: (HR=0.87, 95% CI 0.75-1.0, p=0.05)  Phase III vs Phase I: (HR=0.28, 95% CI 0.21-0.38, p<0.001) | Individual institution vs industry funding: (HR=1.54, 95% CI 1.33-1.78, p<0.0001) | Didn't investigate | Didn't investigate | Didn't investigate | Phase, funding source |
| Stensland et al. 2021 ^47^ | Logistic regression | None significant | Didn't investigate | None significant | Didn't investigate/  Didn't investigate | None significant | None significant | Didn't investigate | None significant | Health service area level prostate cancer incidence per 100 cases: (OR=0.98, 95% CI 0.96-0.99, p=0.03) | Number of centers, phase, funding source, intervention type, anticipated accrual, prostate cancer incidence |
| Dufetelle et al. 2018 ^49^ | Logistic regression | Didn't investigate | Not significant in univariate analysis, excluded from multivariate | Didn't investigate | Completion: Single blind vs open label: (OR=1.42, 95% CI 1.28-1.57, p<0.001)  Completion: Double blind vs open label: (OR=1.17, 95% CI 1.08-1.27, p<0.001)/  Didn’t investigate | Not significant in univariate analysis, excluded from multivariate | Completion: Other funding vs industry: (OR=0.54, 95% CI 0.51-0.58, p<0.001) | Completion: Prevention vs treatment: (OR=1.69, 95% CI 1.52-1.87, p<0.001  Other purpose vs treatment: (OR=1.65, 95% CI 1.46-1.87, p<0.001) | Completion: Device/procedure vs drug/biological: (OR=0.87, 95% CI 0.79-0.95, p=0.002)  Other intervention vs drug/biological: (OR=2.01, 95% CI 1.82-2.22, p<0.001) | Completion: Children vs adults: (OR=1.20, 95% CI 1.05-1.37, p=0.006)  Mixed ages vs adults: (OR=1.19, 95% CI 1.07-1.33, p=0.002);Placebo vs no placebo: (OR=0.90, 95% CI 0.83-0.98, p=0.010)  Cross-over vs parallel: (OR=1.96, 95% CI 1.76-2.17, p<0.001) | Blinding, funding source, primary purpose, intervention type, placebo use, assignment model, age group |
| Hartwell et al. 2022 ^50^ | Logistic regression | Completion: Per participant: (OR=1.04, 95% CI 1.00-1.08, p<0.05) | Didn't investigate | Didn't investigate | Didn't investigate/  None significant | None significant | None significant | Didn't investigate | Didn't investigate | Didn't investigate | Sample size, randomization, phase, funding source, age group |
| van den Bogert et al. 2017 ^52^ | Poisson regression | None significant | Didn't investigate | None significant | Didn't investigate/  None significant | None significant | None significant | Didn't investigate | Didn't investigate | Oncology vs other disease areas: (IRR=1.7, 95% CI 1.1-2.7, p<0.05) | Sample size, randomization, phase, funding source, number and location of centers, specialty |
| Hu et al. 2023 ^53^ | Cox regression | 1-99 vs ≥100: (HR=5.29, 95% CI 2.97-9.42, p<0.001) | United States vs non-United States: (HR=2.64, 95% CI 1.57-4.44, p<0.001) | Didn't investigate | None significant/  None significant | None significant | Industry funding vs other funding: (HR=2.37, 95% CI 1.33-4.25, p=0.004) | None significant | None significant | Didn't investigate | Sample size, location, blinding, randomization, phase, funding source, primary purpose, intervention type |
| Wang et al. 2022 ^54^ | Cox regression | 0-100 vs >100: (HR=4.93, 95% CI 2.02-12.01, p<0.001) | None significant | Didn't investigate | None significant/  None significant | Didn't investigate | Industry vs Other funding: (HR=3.09, 95% CI 1.15-8.34, p=0.026) | None significant | None significant | Didn't investigate | Sample size, location, blinding, randomization, funding source, primary purpose, intervention type |
| Binko et al. 2023 ^55^ | Logistic regression | Completion: ≥100 vs 0-99 : (OR=7.89, 95% CI 1.63-38.3, p=0.01) | Didn't investigate | None significant | None significant/  Didn't investigate | Didn't investigate | None significant | Didn't investigate | None significant | Didn't investigate | Sample size, number of centers, blinding, funding source, intervention type, recruitment national or international |
| Chen et al. 2024 ^56^ | Logistic regression | Completion: Per participant: (OR 1.001, 95% CI 1.000202-1.00218, p=0.02) | Completion: Asia vs North America: (OR 11.57, 95% CI 2.59-51.73, p=0.001)  Europe vs North America: (OR 4.44, 95% CI 1.91-10.3, p=0<0.001) | Not significant in univariate analysis, excluded from multivariate | Didn't investigate/  Didn't investigate | Didn't investigate | Didn't investigate | Completion: Predictive role of AI vs other roles: (OR 4.55, 95% CI 1.44-14.36, p=0.01) | Didn't investigate | Didn't investigate | Sample size, location, primary purpose |
| Alhajahjeh et al. 2024 ^59^ | Logistic regression | <50 vs ≥50: (OR=5.26, 95% CI 3.85-7.69, p<0.001) | Didn't investigate | Single center vs multicenter: (OR=2.11, 95% CI 1.59-2.81, p<0.001) | None significant/  Didn't investigate | None significant | University vs industry funding: (OR=2.20, 95% CI 1.45-3.32, p<0.001) | Didn't investigate | Other vs Device (OR 0.27, 95% CI 0.06-0.99, p=0.049) | Didn't investigate | Sample size, number of centers, blinding, phase, funding source, intervention type, number of agents |
| Buergy et al. 2020 ^60^ | GLM | Didn't investigate | US vs non-US: (OR=2.91, 95% CI 1.90-4.44, p<0.001) | None significant | Didn't investigate/  Didn't investigate | Didn't investigate | None significant | Symptom-control in curative setting vs not: (OR=0.25, 95% CI 0.11-0.56, p=0.001) | Systemic drug intervention vs not: (OR=2.78, 95% CI 1.19-6.48, p=0.018) | Drug intervention x Hematologic trials: (OR=0.244, 95% CI 0.062-0.951, p=0.042)  Multiple interventions x Life-extending interventions: (OR=0.165, 95% CI 0.030-0.897, p=0.037) | Location, funding source, primary purpose, intervention type, cancer type |
| Lee et al. 2020 ^61^ | Cox regression | Didn't investigate | None significant | None significant | Didn't investigate/  Didn't investigate | Didn't investigate | Completion: Industry funded vs not industry funded: (HR=0.66, 95% CI 0.49-0.89, p<0.01) | Completion: Pharmacology vs Product studies: (HR=1.69, 95% CI 1.10-2.59, p=0.02) | Didn't investigate | Didn't investigate | Location, number of centers, funding source, primary purpose |
| Mishra et al. 2024 ^62^ | Logistic regression | Didn't investigate | Didn't investigate | Didn't investigate | None significant/  Didn't investigate | None significant | Other (academic/individual/community) vs industry funding: (OR=6.095, 95% CI 1.078-34.478, p=0.041)  NIH vs industry funding: (OR=0.321, 95% CI 0.108-0.952, p=0.040) | Didn't investigate | None significant | Didn't investigate | Blinding, phase, funding source, intervention type, allocation model, condition |
| Bao et al. 2023 ^63^ | Cox regression | ≥50 vs <50: (HR=0.30, 95% CI: 0.12-0.76, p=0.011) | Non-US vs US location: (HR=0.33, 95% CI: 0.13-0.85, p=0.022) | Didn't investigate | Didn't investigate/  Didn't investigate | Didn't investigate | Not significant in univariate analysis, excluded from multivariate | Didn't investigate | Didn't investigate | Didn't investigate | Sample size, location |
| Roddick et al. 2017 ^65^ | Logistic regression | <20 vs >500: (OR=117.69, 95% CI 24.80-558.40, p<0.001) | Not significant in univariate analysis, excluded from multivariate | Didn't investigate | Double blind vs open label: (OR=3.51, 95% CI 1.36-9.06, p=0.009)/  None significant | Phase 3 vs Phase 2: (OR=5.00, 95% CI 1.86-13.42, p=0.002)  Phase 4 vs Phase 2: (OR=5.67, 95% CI 2.26-14.25, p<0.001) | Industry funding vs academic institutions: (OR=3.89, 95% CI 1.54-9.83, p=0.004)  Industry with other sources vs academic institutions: (OR=5.81, 95% CI 1.95-17.27, p=0.002) | None significant | Device therapies vs pharmacological: (OR=2.70, 95% CI 1.05-6.98, p=0.040)  Other interventions vs pharmacological: (OR=0.15, 95% CI 0.02-0.97, p=0.047) | Heart failure vs coronary artery disease: (OR=2.83, 95% CI 1.23-6.51, p=0.015)  Atrial fibrillation vs coronary artery disease: (OR=3.10, 95% CI 1.21-7.96, p=0.019) | Sample size, blinding, randomization, phase, funding source, primary purpose, intervention type, disease |
| Tanemura et al. 2021 ^67^ | Logistic regression | Didn't investigate | Didn't investigate | Didn't investigate | Didn't investigate/  Didn't investigate | Didn't investigate | Didn't investigate | Didn't investigate | Didn't investigate | Per one country increment: (OR=1.3, 95% CI 1.1-1.5) | Number of countries, trial part of agreed pediatric investigation plan, rare disease, inclusion of newborns/infants/toddlers |
| Shah et al. 2022 ^68^ | Logistic regression | Didn't investigate | Didn't investigate | Didn't investigate | Didn't investigate/  Didn't investigate | None significant | None significant | Diagnostic vs Treatment: (OR=2.952, 95% CI 1.530-5.698, p=0.001) | None significant | Didn't investigate | Phase, funding source, primary purpose, intervention type, allocation model |
| Abdulelah et al. 2024 ^69^ | Logistic regression | Per participant (OR=0.981, p<0.05) | Didn't investigate | None significant | Didn't investigate/  Didn't investigate | Didn't investigate | None significant | Didn't investigate | Didn't investigate | Didn't investigate | Sample size, number of centers, funding source, gender |
| Stensland et al. 2021 ^70^ | Cox regression | Anticipated accrual per additional patient: (HR=0.998, 95% CI 0.997-0.999, p=0.002) | Both USA and international sites vs USA only: (HR=0.34, 95% CI 0.19-0.59, p<0.001)  Non-USA only sites vs USA-only: (HR=0.54, 95% CI 0.38-0.77, p<0.001) | Single center vs multicenter: (HR=1.34, 95% CI 1.02-1.77, p=0.04) | Didn't investigate/  Didn't investigate | Phase 3 vs Phase 2: (HR=1.64, 95% CI 1.01-2.66, p=0.046) | Didn't investigate | Didn't investigate | Drug vs non-drug trials: (HR=1.72, 95% CI 1.04-2.82, p=0.03) | Didn't investigate | Location, number of centers, phase, intervention type, target sample size, trial start year, disease |
| Hecht et al. 2023 ^71^ | Cox regression | 50-100 vs <50: (HR=0.29, 95% CI 0.25-0.35, p<0.001)  101-250 vs <50: (HR=0.25, 95% CI 0.21-0.30, p<0.001)  251-500 vs <50: (HR=0.30, 95% CI 0.24-0.38, p<0.001)  >500 vs <50: (HR=0.29, 95% CI 0.22-0.39, p<0.001) | Didn't investigate | Didn't investigate | Double masking vs None, open label: (HR=0.85, 95% CI 0.74-0.98, p=0.030)/  Didn't investigate | Phase 2 vs Phase 1: (HR=1.35, 95% CI 1.09-1.69, p=0.010)  Phase 3 vs Phase 1: (HR=1.39, 95% CI 1.09-1.78, p=0.010)  Phase 4 vs Phase 1: (HR=1.44, 95% CI 1.14-1.81, p=0.010)  Not applicable phase vs Phase 1: (HR=1.26, 95% CI 1.01-1.59, p=0.041) | Industry vs Government funding: (HR=1.63, 95% CI 1.18-2.25, p=0.003)  Other funding vs Government: (HR=1.69, 95% CI 1.23-2.33, p=0.001) | None significant | Device vs behavioral intervention: (HR=1.63, 95% CI 1.20-2.21, p=0.002)  Drug vs behavioral intervention: (HR=1.48, 95% CI 1.10-2.02, p=0.013) | Pediatrics vs Trauma: (HR=0.58, 95% CI 0.40-0.86, p=0.007)  No results vs Results reported: (HR=1.92, 95% CI 1.68-2.18, p<0.001)  Women vs Any: (HR=0.59, 95% CI 0.45-0.78, p<0.001)  Crossover vs Parallel assignment: (HR=0.63, 95% CI 0.47-0.82, p<0.001)  Single group vs Parallel assignment: (HR=0.69, 95% CI 0.59-0.81, p<0.001) | Sample size, blinding, phase, funding source, primary purpose, intervention type, subspeciality, results reporting, gender, assignment model |
| Steinberg et al. 2024 ^72^ | Logistic regression | Didn't investigate | Didn't investigate | Didn't investigate | Didn't investigate/  Didn't investigate | Didn't investigate | Didn't investigate | Didn't investigate | Didn't investigate | Female vs male PI: (OR=0.58, 95% CI 0.44-0.77, p<0.05) | Sample size, number of centers, blinding, phase, funding source, primary purpose, subspecialty, number of arms, DMC, multiple PI, year of trial registration |
| Steinberg et al. 2022 ^74^ | Cox regression | 0-9 vs 100-599: (HR=39.44, 95% CI 30.43-51.12, p<0.0001)  10-49 vs 100-599: (HR=4.85, 95% CI 3.72-6.32, p<0.0001)  50-99 vs 100-599: (HR=1.98, 95% CI 1.45-2.71, p<0.0001) | Only low and middle income countries vs Includes high income countries: (HR=0.69, 95% CI 0.49-0.96, p=0.03) | Multicenter vs single center: (HR=1.31, 95% CI 1.09-1.57, p=0.004) | None significant/  None significant | Phase 1 vs Phase 2/3-3: (HR=0.60, 95% CI 0.43-0.82, p=0.001)  Not Applicable phase vs Phase 2/3-3: (HR=0.69, 95% CI 0.54-0.89, p=0.004) | Academic funding vs industry funding: (HR=0.63, 95% CI 0.53-0.75, p<0.0001)  Government funding vs industry: (HR=0.49, 95% CI 0.37-0.66, p<0.0001) | None significant | Didn't investigate | Reproductive endocrinology vs gynecologic oncology: (HR=2.08, 95% CI 1.59-2.71, p<0.0001)  Family planning vs gynecologic oncology: (HR=1.55, 95% CI 1.06-2.25, p=0.02)  Multi Arm vs Single Arm: (HR=1.58, 95% CI 1.16-2.16, p=0.004)  First Submission Year: (HR=1.03, 95% CI 1.00-1.06, p=0.02) | Sample size, number of centers, blinding, randomization, phase, funding source, primary purpose,subspecialty, location (income), number of arms, first submission year, data monitoring committee |
| Jacobsen et al. 2023 ^75^ | Logistic regression | None significant | Didn't investigate | Didn't investigate | Didn't investigate/  Didn't investigate | Didn't investigate | None significant | Didn't investigate | Medical device vs pharmaceutical: (OR=2.12, 95% CI 1.05-4.33, p<0.05)  Medical device vs behavioral: (OR=11.61, 95% CI 1.33-101.00, p<0.05 | Didn't investigate | Sample size, funding source, intervention type |
| Johnson et al. 2019 ^76^ | Logistic regression | None significant | Didn't investigate | Didn't investigate | Didn't investigate/  Didn't investigate | Didn't investigate | None significant | Didn't investigate | None significant | Didn't investigate | Sample size, funding source, intervention type |
| Shepard et al. 2023 ^77^ | Logistic regression | ≥80 vs <80: (OR: 0.42, 95% CI 0.2-0.88) | Didn't investigate | Didn't investigate | Didn't investigate/  Didn't investigate | Didn't investigate | None significant | Didn't investigate | None significant | Didn't investigate | Sample size, funding source, intervention type |
| Stensland et al. 2014 ^79^ | Cox regression | Didn't investigate | Non-US vs US trials: (HR=0.65, 95% CI 0.55-0.77, p<0.05)  Both US and non-US vs US trials: (HR=0.67, 95% CI 0.51-0.88, p<0.05) | Single center vs multicenter: (HR=1.93, 95% CI 1.64-2.27, p<0.05) | Didn't investigate/  Didn't investigate | Phase 1/2 vs Phase 3: (HR=1.48, 95% CI 1.12-1.94, p<0.05)  Phase 2 vs Phase 3: (HR=1.29, 95% CI 1.02-1.62, p<0.05) | Industry vs federal funding: (HR=1.97, 95% CI 1.57-2.48, p<0.05) | Didn't investigate | Behavioral vs Drug/supplement: (HR=0.24, 95% CI 0.06-0.96, p<0.05) | Lung vs Hematologic cancer: (HR=1.32, 95% CI 1.04-1.69, p<0.05) | Location, number of centers, phase, funding source, intervention type, disease |
| Bandari et al. 2020 ^80^ | Logistic regression | Didn't investigate | Didn't investigate | Didn't investigate | Didn't investigate/  None significant | None significant | Combined industry/government/grants vs Industry funding: (OR=3.13, 95% CI 2.21-4.48, p<0.05 | Didn't investigate | Device vs Drug trials: (OR=1.64, 95% CI 1.00-2.70, p=0.05) | Oncology vs General subspecialty: (OR=2.25, 95% CI 1.60-3.18, p<0.05)  Infertility/andrology vs General subspecialty: (OR=4.99, 95% CI 1.60-17.61, p<0.05) | Randomization, phase, funding source, intervention type, subspecialty, American Urological Association section |
| Spinosa et al. 2021 ^86^ | Logistic regression | Didn't investigate | US-only (OR=1.35 95% CI=1.02-1.77) | 2-5 sites vs single (OR=0.66 95% CI=0.47-0.93) | Didn't investigate/  Didn't investigate | Phase 1/2 or 2 trials vs early phase 1 or 1 (OR=1.54 95% CI=1.13-2.11) | Industry funded (OR=1.45 95% CI=1.03-2.03) | None significant | None significant | Didn't investigate | Location, number of centers, phase, funding source, primary purpose, intervention type, cancer type, primary outcomes, NCI involvement |

**eAppendix 5 Significant predictors of failure in multivariable regression analyses**

The most convincing predictor of trial failure was smaller sample size (i.e. enrollment). It was explored in 25 studies and found to be a statistically significant predictor of failure in multivariable analyses in 18 studies ^9, 18, 23, 29, 37, 38, 50, 53, 54, 55, 56, 59, 63, 65, 69, 71, 79, 85^. On the other hand, eight studies explored planned sample size/anticipated accrual as opposed to actual sample size ^15, 24, 25, 31, 40, 41, 47, 70^, and it was found not to be a statistically significant predictor in five studies ^5, 24, 31, 40, 47^, protective factor in two studies ^25, 70^, and predictive of failure in one study ^41^.

Funding source was the most examined predictor and it was explored in 47 studies ^9, 11, 12, 13, 14, 15, 16, 18, 19, 22, 23, 25, 26, 28, 29, 30, 32, 34, 38, 39, 40, 41, 43, 44, 47, 49, 50, 52, 53, 54, 55, 59, 60, 61, 62, 63, 65, 68, 69, 71, 76, 79, 83, 84, 85, 87, 80^, and was a significant predictor in 24 studies ^9, 11, 12, 13, 22, 25, 28, 29, 30, 38, 39, 44, 49, 53, 54, 59, 61, 62, 65, 71, 76, 79, 87, 80^. Industry funding was found to be predictive of failure in 8 out of 9 cases when compared with government funding ^9, 27, 29, 38, 62, 71, 79, 87^ , and protective in study ^28^; 6 out of 7 cases when compared with academic funding ^9, 25, 29, 38, 65, 79^, and protective in study ^59^, all three times when compared with non-industry funding ^22, 61, 76^, and both times when compared with local funding ^12, 13^. On the other hand, it was found to be protective of failure in one study each when compared with healthcare center funding^39^ and with individual funding^44^. When compared with other funding, it was found to be protective in three studies ^29, 53, 54^, and predictive of failure in five studies ^11, 30, 39, 49, 62^. However, the definitions of other funding varied across studies.

The geographic location of the trial as a predictor was investigated in 18 studies, with US-based trials being statistically significant predictors of failure compared with non-US based or combined in 10 studies ^15, 30, 43, 53, 56, 60, 63, 70, 76, 87^, and 8 studies finding no statistically significant difference ^9, 11, 32, 34, 49, 54, 61, 65^. Additional four studies investigated location in terms of income as a predictor, with all four ^29, 38, 41, 79^ finding that trials performed in higher income countries are more likely to prematurely terminate. The number of participating countries was investigated in only three studies, with two finding no significance ^15, 41^, and one finding increased number as a risk factor ^67^.

On the other hand, the number of participating centers as a predictor of premature termination was investigated in 21 studies and yielded inconsistent results ^9, 15, 23, 24, 29, 32, 34, 38, 40, 47, 52, 55, 56, 59, 60, 61, 69, 70, 76, 79, 87^. Namely, 13 studies found no statistically significant differences, four studies reported single center trials being predictive of failure compared to multicenter trials ^59, 70, 76, 87^, while three studies found the opposite ^9, 38, 79^, and one study observed a protective effect of a higher number of centers in increments of 10 centers ^15^.

The trial phase was among the most examined factors, with 33 studies exploring it ^9, 12, 13, 14, 16, 19, 22, 23, 29, 30, 31, 32, 34, 38, 39, 41, 43, 44, 47, 49, 50, 52, 53, 59, 62, 65, 68, 70, 71, 76, 79, 87, 80^. However, only 14 studies found it to be a significant predictor ^9, 13, 23, 29, 30, 38, 43, 44, 65, 70, 71, 76, 79, 87^, and the findings in regards which phase was a risk factor were inconsistent. The exception is the finding in 8 studies that phase 1 studies were of less risk to failure compared to other phase studies ^9, 29, 30, 38, 43, 71, 76, 79^ while two studies showed the opposite ^44, 87^.

Some methodological factors were rarely found to be significant predictors. Blinding was investigated in 23 studies ^9, 12, 13, 16, 19, 23, 25, 28, 29, 32, 34, 38, 39, 41, 49, 53, 54, 55, 59, 62, 65, 71, 79^, with 17 studies finding no association, three studies finding it predictive of failure ^12, 38, 65^ and three finding it protective of failure ^19, 49, 71^. Similarly, randomization was explored in 16 studies ^9, 12, 13, 16, 22, 29, 30, 38, 41, 50, 52, 53, 54, 65, 79, 80^, and found to be significant only in two studies, both finding it predictive of failure ^29, 38^. The presence of data monitoring committee was found to be predictive of failure ^29^, protective of failure ^38^, and non-significant ^79^ in one study each.

1. At least one US site
2. 662 shoulder-related + 126 elbow-related
3. Failure proportions per group: Shoulder - 8%, Elbow - 13%
4. Except for Terminated for "Good" reasons
5. “Good” terminated - terminations because of safety or efficacy reasons
6. 10.2% both terminated for good reasons and bad reasons
7. For reasons other than interim results or toxicity
8. For interim results or toxicity issues
9. Pharmaceutical interventions
10. At least one comparator group
11. Defined as trials that explicitly reported discontinuation in a journal publication/survey query OR achieved <90% of target sample size with reported recruitment problems
12. 82,719 nonsurgical + 5,779 surgical
13. US-registered
14. For surgical trials; nonsurgical 11.07%. Absolute numbers not provided, the overall failure rate cannot be calculated
15. Effect and safety studies
16. Preplanned sample size ≥100
17. Defined as not meeting preplanned sample size
18. Involve external beam radiation, actively receiving radiotherapy
19. Data not provided
20. Trials using indices or composite measures
21. Rare, non-infectious diseases
22. Defined as trials that explicitly reported discontinuation in a trial registry/ publication/communication OR achieved <90% of

    target sample size
23. Never started and Ongoing excluded
24. At least one drug intervention
25. Belonging to one of top 25 countries (≥1000 trials)
26. For period 2002-2006. No data is provided for 2007-2008. The predictors were estimated implementing a 3:1 matched

    case-control design using the 84 terminated trials from the period 2002-2008.
27. Defined as trials that ended prior to the last treatment of the last patient according to the last version of the protocol
28. 2002-2006 data
29. US-based
30. Involving incision to head or spine and/or had neurosurgeon as principal investigator
31. Failure proportions per group: Adult – 10.5%, Pediatric - 8.5%
32. The authors stated in the discussion that the failed proportion is 4.3%. However, this information isn’t presented in the

    results, and it doesn't match the numbers provided
33. ToetsingOnline database contains all IRB-reviewed clinical trials in the Netherlands
34. AI as an intervention
35. Trials suspended for ≥ 1.5 years
36. Microbubble-based contrast agents
37. The trial design was case-control, for every failed trial a completed control was found
38. Trials that have been halted by the sponsor for different reasons but that may start again
39. Trials that have been halted by the drug administrative department but it will no longer start again
40. Countries in EU/EEA
41. A trial for which a negative ethics committee opinion was issued in any of EU member states and that could therefore not be

    initiated in that state
42. A trial that has been temporarily interrupted
43. A trial that ended before the completion of the all procedures described in the protocol.
44. Good terminated - terminations because of toxicity/adverse events, interim results, or trial no longer needed
45. 14.7% both terminated for good reasons and bad reasons
46. “Trials were considered prematurely discontinued if the accrual was closed before recruiting the target sample size. For the

    trials without available data on actual enrollment, a standardized e-mail was sent to the contact e-mail address on

    ClinicalTrials.gov to confirm the discontinuation status. The trials were also deemed as discontinued if the trial investigators

    could not be contacted.”
47. Conference abstract
48. Definition unclear
49. "Discontinued early" is stated alongside withdrawn and suspended
50. Possibly all categories: " All heart failure clinical trials registered on CTG from September 27, 2007 to March 31, 2022 were

    identified"
51. Possibly only completed: "...to identify all completed and discontinued clinical trials in cardiovascular medicine…"
52. Possibly only completed: "...to identify all completed and discontinued heart failure clinical trials…"
53. Most likely: "Terminated before meeting target enrollment”
54. Most likely: “The trials were categorized into two groups: terminated and completed”
55. Abstract of an unavailable paper
